# Transdiagnostic metabolic architecture across major depressive disorder and schizophrenia: a NIMETOX systems-biology approach

**DOI:** 10.64898/2026.09.28.26364141

**Authors:** Yueyang Luo, Hongzhou Wu, Chenghui Yang, Tangcong Chen, Abbas F Almulla, Yingqian Zhang, Michael Maes

**Author notes:** Corresponding Authors: Prof. Dr. Michael Maes, M.D., Ph.D., Dr. HC, and Dr. Yingqian Zhang Sichuan Provincial Center for Mental Health, Sichuan Provincial People’s Hospital, School of Medicine, University of Electronic Science and Technology of China, Chengdu 610072, China. Co-Joint first authors (Yueyang Luo, Hongzhou Wu). Prof. Dr. Michael Maes, M.D., Ph.D. https://scholar.google.co.th/citations?user=1wzMZ7UAAAAJ&hl=th&oi=ao, Highly cited author: 2003-2023 (ISI, Clarivate), Scholar GPS: Worldwide #1 in molecular neuroscience; #1/4 in pathophysiology Expert worldwide medical expertise ranking, Expertscape (December 2022), worldwide: #1 in CFS, #1 in oxidative stress, #1 in encephalomyelitis, #1 in nitrosative stress, #1 in nitrosation, #1 in tryptophan, #1 in aromatic amino acids, #1 in stress (physiological), #1 in neuroimmune; #2 in bacterial translocation; 3 in inflammation, #4-5: in depression, fatigue, and psychiatry.

## Abstract

**Background:** Major depressive disorder (MDD) is conceptualised within a neuroimmune–metabolic–oxidative (NIMETOX) framework. From a systems-biology perspective, related metabolic–oxidative disturbances may extend across diagnostic boundaries, including schizophrenia (SCZ), but their relationships with childhood adversity and dimensional psychopathology remain unclear.

**Objectives:** To characterize shared and divergent serum metabolic signatures across MDD and SCZ and their associations with childhood adversity, resilience, and dimensional clinical phenotypes.

**Methods:** We studied 89 participants (26 HC, 34 MDD, 29 SCZ). Untargeted serum metabolomics yielded 1,419 metabolites, of which 38 selected markers were integrated into 11 metabolic modules. Group comparisons, nested cross-validated OPLS-DA, and multiple regression analyses were performed.

**Results:** Compared with HC, MDD and SCZ exhibited reduced mitochondrial lipid bioenergetics and metabolic resilience and increased sphingolipid stress signalling, oxidative lipid damage, and renin–angiotensin–aldosterone system stress. PUFA membrane remodelling increased progressively from HC to SCZ to MDD, whereas antioxidants were selectively reduced in MDD. An integrated OPLS-DA discriminated HC from MDD+SCZ (cross-validated AUC=0.946). Across dimensional models, mitochondrial lipid bioenergetics, PUFA remodelling, sphingolipid stress, oxidative lipid damage, and metabolic resilience explained substantial variance in mood, positive and negative psychotic symptoms, resilience, and suicidal behaviors. A terpenoid exposure–biotransformation module was strongly associated with psychosis and suicidal behaviors.

**Conclusion:** MDD and SCZ share an interconnected NIMETOX metabolic architecture while clinical dimensions show partly distinct psychosocial–metabolic configurations. These findings support a transdiagnostic NIMETOX systems-biology model linking NIMETOX dysfunction with dimensional psychopathology and provide a framework for future stratification and biomarker validation.

## Introduction

Major depressive disorder (MDD) and schizophrenia (SCZ) are clinically distinct psychiatric disorders, yet accumulating molecular evidence indicates substantial overlap in their peripheral metabolic abnormalities. In MDD, these disturbances have increasingly been conceptualised within the neuroimmune–metabolic–oxidative (NIMETOX) framework, which integrates immune dysregulation with disturbances in energy metabolism, lipid biology, oxidative stress, and related cellular signalling [1, 2]. There is also evidence that NIMETOX pathways play a key role in SCZ [3, 4]. Comparative lipidomic and metabolomic studies likewise indicate extensive molecular overlap between MDD and SCZ, accompanied by a smaller subset of disorder-dependent features, including sphingolipid-related and other lipid differences [5–7]. Together, these findings suggest that MDD and SCZ may share a transdiagnostic metabolic background on which selective disorder-related variation is superimposed.

Within this broader metabolic context, lipid–bioenergetic–redox dysregulation may represent a particularly informative biological layer [1, 4]. Polyunsaturated fatty acid (PUFA) incorporation and phospholipid remodelling influence membrane phospholipid composition and lipid-mediated signalling, whereas sphingolipid pathways intersect with mitochondrial bioenergetics and redox homeostasis [8, 9]. In MDD, altered circulating PUFA composition and broader phospholipid abnormalities have been reported [10, 11]. More recent deep metabolomic phenotyping further suggests coordinated disturbances involving lipid remodelling, fatty-acid metabolism, mitochondrial redox balance, ether-lipid metabolism, and antioxidant capacity [12].

Oxidative abnormalities, including lipid peroxidation and impaired antioxidant defence, have likewise been documented in SCZ [13, 14]. Together, these observations support examining lipid, bioenergetic, and redox systems as an interconnected metabolic architecture rather than as isolated pathways.

Beyond diagnosis, metabolic heterogeneity may also reflect psychosocial history, clinical phenotype, and exposure-sensitive variation. Childhood trauma is a transdiagnostic vulnerability factor for psychiatric illness [15]. Such adversity may become biologically embedded through persistent neuroendocrine, immune, and metabolic alterations [16, 17]. Childhood adversity has been associated with altered circulating metabolic profiles [18, 19] and with alterations in the renin–angiotensin–aldosterone system (RAAS) [20]. At the clinical level, peripheral metabolic and redox abnormalities have been linked to depressive symptom dimensions, psychotic symptoms, and suicidality [21–23]. Emerging evidence also suggests that psychological resilience may have peripheral metabolic correlates [24]. Finally, circulating metabolite profiles may reflect dietary, behavioural, and environmental influences, positioning the metabolome at the interface between endogenous physiology and the broader exposome [25, 26].

Despite these advances, the relationships among these domains remain poorly integrated. It remains unclear to what extent MDD and SCZ share a serum metabolic architecture while retaining selective diagnostic differences, or how childhood adversity relates to that architecture. Whether distinct metabolic configurations map onto mood, behavioural, psychotic, suicidal, and adaptive dimensions is uncertain.

Existing studies have generally examined these domains separately rather than within a single transdiagnostic serum framework. Resolving these relationships may help distinguish shared metabolic disturbances from diagnostic, clinical, psychosocial, and exposure-related heterogeneity.

Accordingly, the present study used untargeted serum metabolomics to characterize the transdiagnostic metabolic architecture across MDD or SCZ. Specifically, we aimed to (1) identify shared and diagnostically divergent patterns across metabolic modules and evaluate the multivariate metabolic profile discriminating between controls and participants with mental disorders (MDD and SCZ combined); (2) examine how these metabolic features relate to childhood adversity and dimensional outcomes, including overall severity of mood (OSOM), positive and negative symptoms, overall severity of psychosis (OSOP), suicidal ideation and behaviors, overall severity of illness (OSOI), and psychological resilience. We hypothesized that MDD and SCZ would exhibit a shared bioenergetic–lipid–oxidative metabolic background with selective diagnostic divergence, and that distinct metabolic–psychosocial configurations would be associated with specific symptom and resilience dimensions.

## Methods

### Participants and clinical assessments

The study included 89 participants: 26 healthy controls (HC), 34 MDD, and 29 SCZ, aged 18–65 years. MDD and SCZ participants were recruited from the Sichuan Mental Health Center, Sichuan Provincial People’s Hospital, Chengdu, China. HC were recruited contemporaneously from hospital staff and their social contacts and were matched to the clinical groups for age, sex, body mass index (BMI), and educational attainment. BMI was calculated as body weight in kilograms divided by height in meters squared (kg/m²). Diagnoses were established by a senior psychiatrist according to DSM-5 criteria and further assessed using the Mini-International Neuropsychiatric Interview (M.I.N.I. 6.0) [27, 28]. Current psychotropic medication use was recorded at assessment. Detailed eligibility criteria are provided in the Electronic Supplementary File (ESF), Method S1.

Clinical assessments included the Columbia-Suicide Severity Rating Scale (C-SSRS), Positive and Negative Syndrome Scale (PANSS), Beck Depression Inventory-II (BDI-II), State-Trait Anxiety Inventory, state version (STAI-state), Connor–Davidson Resilience Scale (CD-RISC), and Childhood Trauma Questionnaire–Short Form (CTQ-SF) [29–34]. Derived clinical dimensions comprised current suicidal ideation, total suicidal behaviour, positive and negative symptoms, OSOM, OSOP, OSOI, and psychological resilience. Detailed scoring and construction of the derived clinical variables are provided in ESF, Method S1.

The study was approved by the Ethics Committee of the University of Electronic Science and Technology of China (IRB No. 30850). All participants provided written informed consent, and all procedures were conducted in accordance with the Declaration of Helsinki.

### Blood sampling and biochemical measures

Fasting venous blood was collected between 07:00 and 08:00 h, centrifuged, and serum aliquots were stored at −80 °C until analysis. Albumin, transferrin, triglycerides (TG), total cholesterol (TC), LDL-C, HDL-C, apolipoprotein A1 (ApoA1), and free cholesterol (FC) were measured using established assays on an ADVIA 2400 automated biochemical analyser. Derived variables included VLDL-C, the LDL-C/HDL-C ratio, the atherogenic index of plasma [AIP = z(TG) − z(HDL-C)] [35], and an antioxidant-defence composite z[z(albumin) + z(HDL-C) + z(ApoA1)]. Detailed assay procedures and derivation of the biochemical variables are provided in ESF, Method S2.

### Untargeted serum metabolomics

Untargeted serum metabolomic profiling was performed using an ACQUITY UPLC system (Waters) coupled to a Q-Exactive Plus high-resolution mass spectrometer (Thermo Fisher Scientific). Data were acquired separately in positive- and negative-ion modes over m/z 80–1200, with pooled quality-control samples interspersed throughout the analytical sequence. Raw files were converted to mzML format and processed in Progenesis QI version 2.4 for preprocessing, metabolite annotation, and quality-control filtering. Features with an annotation score >0.5 and QC coefficient of variation <0.30 were retained. After merging the positive- and negative-ion datasets, 1,419 annotated metabolite features were available for subsequent analyses. Detailed procedures are provided in ESF, Method S3.

### Metabolite annotation screening and feature selection

The analytical workflow is summarised in **Fig. 1**. The 1,419 annotated metabolite features underwent a study-specific, rule-based screen of Human Metabolome Database (HMDB) annotations, yielding an endogenous-enriched candidate pool of 540 metabolites [36]. Detailed annotation screening, supervised feature selection, and stability selection are described in ESF, Method S4.

**Figure 1.**
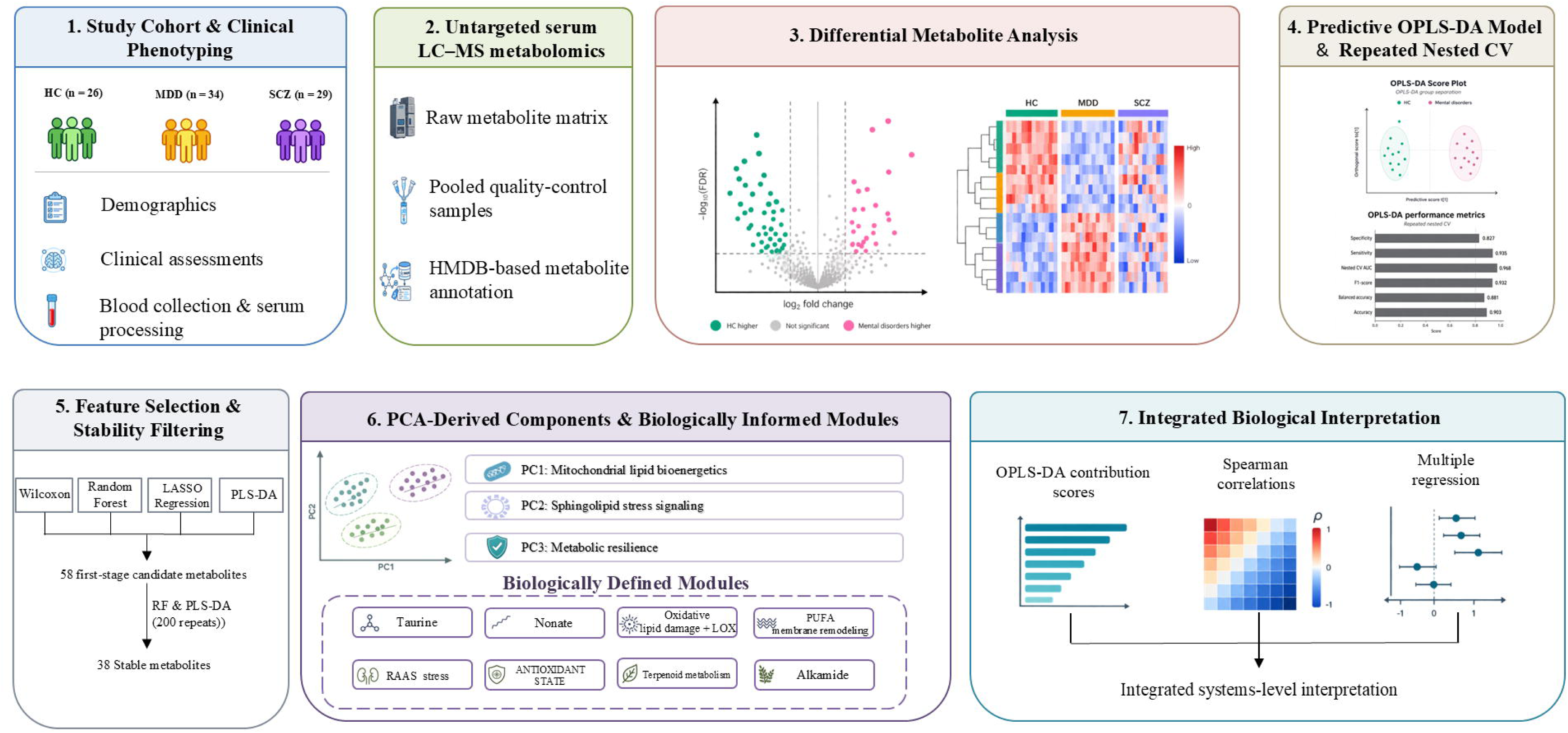
Study design and analytical workflow. The workflow summarises cohort characterisation and clinical phenotyping, untargeted serum LC–MS metabolomics, differential-abundance analysis, metabolite feature selection and stability filtering, predictive OPLS-DA modelling with repeated nested cross-validation, construction of metabolic modules, and integrative systems-level interpretation.

### Construction of metabolic modules

The 38 retained metabolites were used to construct 11 metabolic modules, comprising three Principal component analysis (PCA)-derived components and eight non-PCA modules. Detailed construction procedures are provided in ESF, Method S5.

### Statistical analyses

Continuous variables were compared across HC, MDD, and SCZ using Kruskal–Wallis tests followed, where appropriate, by tie-corrected Dunn tests with Holm adjustment. Categorical variables were assessed using Pearson χ² tests followed by two-sided Fisher’s exact tests with Holm adjustment. Differential abundance across the 1,419 annotated metabolite features was assessed using Linear Models for Microarray Data (limma) on normalised log2 abundances. Pooled MDD+SCZ versus HC was the primary contrast, with diagnosis-specific contrasts examined secondarily. Significance required a Benjamini–Hochberg false discovery rate (FDR) <0.05 and an absolute fold change ≥1.5.

OPLS-DA was used to discriminate HC from pooled MDD+SCZ using the complete metabolomic profile and, separately, an integrated set of metabolic modules, biochemical variables, and childhood-adversity variables. Model significance was assessed using 1,000 label permutations. Predictive performance of both the full-metabolome and integrated OPLS-DA models was assessed using five-fold stratified outer cross-validation repeated 20 times, with five-fold inner cross-validation used to select 0–3 orthogonal components based on mean inner-CV Q². Specificity, sensitivity, AUC, F1-score, balanced accuracy, and overall accuracy were calculated, and 95% intervals were obtained from 2,000 bootstrap resamples of the repeat-level performance estimates. The primary integrated model excluded the two putatively diet-derived modules, terpenoid metabolism and alkamide; alternative configurations varied the inclusion of these two modules and the childhood-adversity domains. The 38-metabolite panel was additionally evaluated using random-forest and elastic-net classifiers trained in the feature-selection subset (n=62) and tested in the locked held-out subset (n=27). Full model specifications and validation procedures are provided in ESF, Method S6.

Spearman rank correlations examined associations between the 11 metabolic modules and demographic, childhood-adversity, and clinical measures, with Benjamini–Hochberg FDR correction applied separately within the demographic/childhood-adversity and clinical panels. Multivariable associations with eight clinical outcomes were examined using stepwise linear regression with repeated cross-validation, bootstrap assessment of coefficient stability, and variance-inflation-factor diagnostics. Detailed association analyses are provided in ESF, Method S7. Sensitivity analyses assessed diagnostic-group differences after adjustment for age, sex, and BMI. Medication sensitivity analyses were restricted to participants with MDD or SCZ and examined current antidepressant, benzodiazepine, mood-stabiliser, and antipsychotic use in separate covariate-adjusted models.

All analyses were performed using R version 4.4.3 and Python version 3.14.4. All tests were two-sided, with P < 0.05 considered statistically significant unless otherwise specified.

## Results

### Demographic and clinical characteristics

The three study groups did not differ significantly in age, sex distribution, BMI, or years of education (**Table 1**). All clinical measures differed significantly across diagnostic groups. SCZ showed higher current suicidal ideation, positive and negative symptoms, and OSOP than both MDD and HC. Total suicidal behaviour was higher in both clinical groups than in HC, with no significant difference between MDD and SCZ. MDD showed higher OSOM than both SCZ and HC. OSOI was higher in both clinical groups than in HC, whereas psychological resilience was highest in HC and lowest in MDD.

**Table 1.** Demographic and clinical characteristics across diagnostic groups.

| Variable | HC (n=26) | MDD (n=34) | SCZ (n=29) | H/$\chi^2$ | P | HC vs MDD | HC vs SCZ | MDD vs SCZ |
| --- | --- | --- | --- | --- | --- | --- | --- | --- |
| Age (years) | 33.9 (14.0) | 28.0 (9.2) | 33.0 (12.6) | 4.32 | 0.116 | — | — | — |
| Male, n (%) | 10 (38.5%) | 12 (35.3%) | 12 (41.4%) | 0.25 | 0.884 | — | — | — |
| BMI (kg/m<sup>2</sup>) | 25.1 (4.9) | 22.7 (3.8) | 21.8 (3.4) | 5.50 | 0.064 | — | — | — |
| Education (years) | 12.8 (3.5) | 13.4 (4.0) | 13.00 (3.1) | 2.15 | 0.341 | — | — | — |
| Current suicidal ideation (z) | -0.90 (0.09) | -0.02 (0.93) | 0.83 (0.81) | 44.53 | <0.001 | <0.001 | <0.001 | <0.001 |
| Total suicidal behaviour (z) | -1.12 (0.18) | 0.29 (0.86) | 0.67 (0.71) | 54.06 | <0.001 | <0.001 | <0.001 | 0.050 |
| Positive symptoms (z) | -1.08 (0.00) | -0.05 (0.64) | 1.02 (0.80) | 66.93 | <0.001 | <0.001 | <0.001 | <0.001 |
| Negative symptoms (z) | -1.07 (0.00) | 0.03 (0.72) | 0.90 (0.88) | 62.08 | <0.001 | <0.001 | <0.001 | 0.002 |
| OSOP (z) | -1.13 (0.00) | -0.01 (0.65) | 1.01 (0.73) | 66.73 | <0.001 | <0.001 | <0.001 | <0.001 |
| OSOM (z) | -1.07 (0.27) | 0.98 (0.56) | -0.33 (0.61) | 64.47 | <0.001 | <0.001 | <0.001 | <0.001 |
| OSOI (z) | -1.39 (0.18) | 0.61 (0.52) | 0.44 (0.65) | 55.96 | <0.001 | <0.001 | <0.001 | 0.244 |
| Psychological resilience (z) | 0.83 (0.63) | -0.79 (0.61) | 0.28 (0.97) | 42.85 | <0.001 | <0.001 | 0.031 | <0.001 |

Data are mean (SD) for continuous variables and n (%) for categorical variables. Group differences were assessed using Kruskal-Wallis tests or Pearson $\chi^2$ tests. Pairwise tests were performed only after a significant omnibus test and were Holm-adjusted across the three comparisons within each variable. All tests were two-sided.
Abbreviations: BMI, body mass index; HC, healthy controls; MDD, major depressive disorder; OSOI, overall severity of illness; OSOM, overall severity of mood; OSOP, overall severity of psychosis; SCZ, schizophrenia; SD, standard deviation; z, standardised score.

Among blood biomarkers, albumin, apolipoprotein A1 (ApoA1), and the antioxidant-defence composite were lower in both clinical groups than in HC. HDL-C was lower in MDD than in HC, whereas the LDL-C/HDL-C ratio and atherogenic index of plasma (AIP) were higher in MDD than in HC. None of these biomarkers differed significantly between MDD and SCZ, and no significant overall group differences were observed for the remaining biochemical variables (**Table 2**).

**Table 2.** Metabolic variables and blood biomarkers across diagnostic groups.

| Variable | HC (n=26)<br>Mean (SD) | MDD (n=34)<br>Mean (SD) | SCZ (n=29)<br>Mean (SD) | H | P | HC vs MDD | HC vs SCZ | MDD vs SCZ |
| --- | --- | --- | --- | --- | --- | --- | --- | --- |
| PC1: Mitochondrial lipid bioenergetics (PC score) | 0.75 (0.82) | -0.36 (0.92) | -0.25 (0.90) | 21.11 | <0.001 | <0.001 | <0.001 | 0.554 |
| PC2: Sphingolipid stress signalling (PC score) | -0.63 (0.90) | 0.38 (0.82) | 0.12 (1.03) | 15.90 | <0.001 | <0.001 | 0.013 | 0.260 |
| PC3: Metabolic resilience (PC score) | 0.53 (0.90) | -0.26 (0.95) | -0.17 (0.99) | 10.97 | 0.004 | 0.006 | 0.015 | 0.725 |
| Taurine (z) | -0.52 (1.17) | 0.27 (0.77) | 0.15 (0.93) | 9.43 | 0.009 | 0.007 | 0.081 | 0.347 |
| Nonate (z) | -0.52 (0.65) | 0.14 (1.13) | 0.31 (0.93) | 11.44 | 0.003 | 0.043 | 0.003 | 0.235 |
| Oxidative lipid damage + LOX (z) | -1.00 (0.78) | 0.50 (0.69) | 0.30 (0.84) | 34.11 | <0.001 | <0.001 | <0.001 | 0.554 |
| PUFA membrane remodelling (z) | -0.93 (0.74) | 0.79 (0.61) | -0.09 (0.81) | 47.40 | <0.001 | <0.001 | 0.002 | <0.001 |
| ANTIOXIDANT STATE (z) | 0.38 (0.69) | -0.52 (0.96) | 0.27 (1.05) | 14.43 | <0.001 | 0.001 | 0.386 | 0.012 |
| RAAS stress (z) | -0.73 (0.91) | 0.52 (0.87) | 0.05 (0.83) | 22.03 | <0.001 | <0.001 | 0.013 | 0.056 |
| Terpenoid metabolism (z) | -1.06 (0.66) | 0.39 (0.70) | 0.49 (0.83) | 40.43 | <0.001 | <0.001 | <0.001 | 0.762 |
| Alkamide (z) | -0.41 (0.85) | 0.16 (0.97) | 0.18 (1.08) | 7.96 | 0.019 | 0.035 | 0.035 | 0.850 |
| Albumin (g/L) | 45.33 (2.18) | 42.33 (3.20) | 43.13 (3.71) | 14.68 | <0.001 | <0.001 | 0.026 | 0.208 |
| Triglycerides (mmol/L) | 1.18 (0.67) | 1.42 (0.76) | 1.36 (0.63) | 2.63 | 0.268 | — | — | — |
| Total cholesterol (mmol/L) | 4.80 (0.98) | 4.87 (0.80) | 4.42 (0.92) | 4.14 | 0.126 | — | — | — |
| HDL-C (mmol/L) | 1.37 (0.27) | 1.14 (0.33) | 1.22 (0.34) | 8.86 | 0.012 | 0.012 | 0.056 | 0.531 |
| LDL-C (mmol/L) | 2.72 (0.89) | 2.98 (0.71) | 2.55 (0.75) | 4.88 | 0.087 | — | — | — |
| LDL-C/HDL-C ratio | 2.10 (0.93) | 2.87 (1.16) | 2.22 (0.86) | 8.99 | 0.011 | 0.014 | 0.478 | 0.062 |
| Apolipoprotein A1 (g/L) | 1.73 (0.27) | 1.37 (0.30) | 1.51 (0.30) | 19.39 | <0.001 | <0.001 | 0.011 | 0.125 |
| Antioxidant-defence composite (z) | 0.66 (0.77) | -0.44 (0.88) | -0.07 (1.03) | 20.84 | <0.001 | <0.001 | 0.003 | 0.204 |
| AIP (z) | -0.38 (0.85) | 0.25 (1.10) | 0.05 (0.93) | 6.57 | 0.037 | 0.038 | 0.124 | 0.567 |

Data are mean (SD). Between-group differences were assessed using Kruskal-Wallis tests. Pairwise Dunn tests were performed only after a significant omnibus test and were Holm-adjusted across the three diagnostic-group comparisons. All tests were two-sided.
Abbreviations: AIP, atherogenic index of plasma; HC, healthy controls; HDL-C, high-density lipoprotein cholesterol; LDL-C, low-density lipoprotein cholesterol; LOX, lipoxygenase; MDD, major depressive disorder; PC, principal component; PUFA, polyunsaturated fatty acid; RAAS, renin-angiotensin-aldosterone system; SCZ, schizophrenia; SD, standard deviation; VLDL-C, very-low-density lipoprotein cholesterol; z, standardised score.

### Serum metabolomic profiles and differential abundance

The global serum metabolomic profile comprising all 1,419 annotated metabolite features is shown in **Fig. 2A**. Considerable inter-individual variation and overlapping patterns were observed across HC, MDD, and SCZ. Differential-abundance analysis comparing pooled MDD+SCZ with HC identified 71 metabolite features that met the predefined criteria of FDR < 0.05 and absolute fold change ≥ 1.5 (**Fig. 3**). Of these, 14 showed higher and 57 lower abundance in pooled MDD+SCZ relative to HC. Diagnosis-specific analyses identified 63 differential features for MDD versus HC and 43 for SCZ versus HC, whereas only two features differed between MDD and SCZ, with one showing higher abundance in each diagnostic group (ESF, Fig. S1).

**Figure 2.**
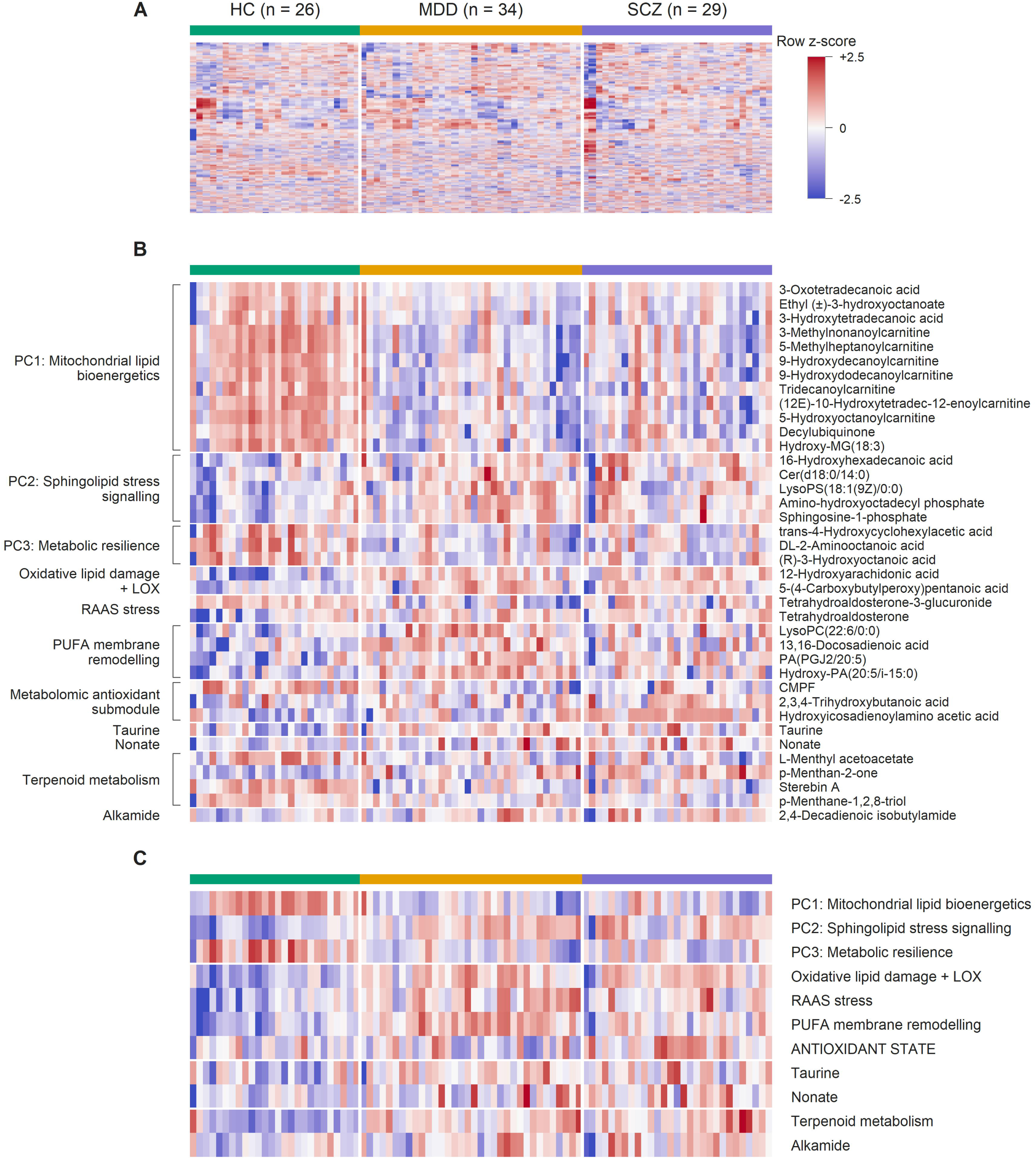
Heatmap profiles of serum metabolites across analytical levels. Heatmaps show the global serum metabolome comprising 1,419 metabolites **(A)**, the 38 stable metabolites retained after feature-selection and stability analyses and organized according to their corresponding biologically informed modules **(B)**, and the resulting 11 biologically informed metabolic modules (C) across controls (HC), major depressive disorder (MDD), and schizophrenia (SCZ). Columns represent the same participants in an identical order across panels, and colors indicate row-wise standardized z-scores capped at −2.5 and +2.5 for visualization; brackets in **(B)** indicate modules represented by three or more constituent metabolites.

**Figure 3.**
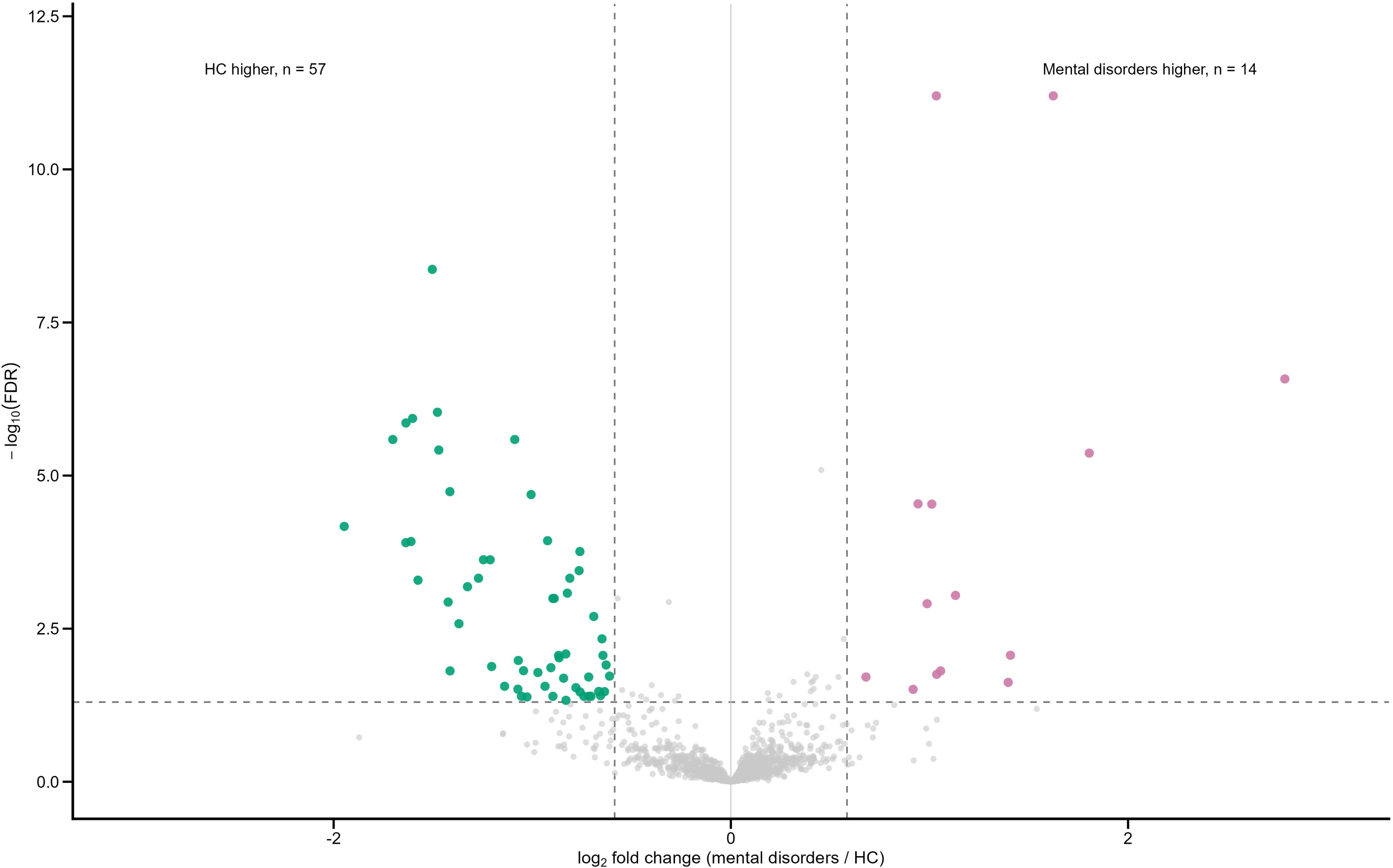
Differential serum metabolite profiles between healthy controls and mental disorders. Volcano plot showing differential serum metabolites between healthy controls (HC) and pooled participants with major depressive disorder (MDD) or schizophrenia (SCZ) (labeled as mental disorders). Each point represents one metabolite. Metabolites meeting the predefined statistical thresholds of false discovery rate (FDR) < 0.05 and absolute fold change ≥ 1.5 are shown in green when higher in HC and in magenta when higher in the pooled clinical group; non-significant metabolites are shown in grey. Vertical dashed lines indicate the fold-change threshold, and the horizontal dashed line indicates the FDR threshold. Numbers indicate metabolites meeting both statistical criteria in each direction.

### OPLS-DA discrimination between HC and mental disorders

OPLS-DA based on all 1,419 annotated metabolite features showed separation between HC and pooled MDD+SCZ (**Fig. 4A)**. Permutation testing based on 1,000 label permutations yielded a Q² intercept of −0.821, with empirical P = 0.001 for both R²Y and Q² (**Fig. 4B**). Repeated nested cross-validation yielded a specificity of 0.887, sensitivity of 0.975, AUC of 0.965, F1-score of 0.964, balanced accuracy of 0.931, and overall accuracy of 0.949 (**Fig. 4C**).

**Figure 4.**
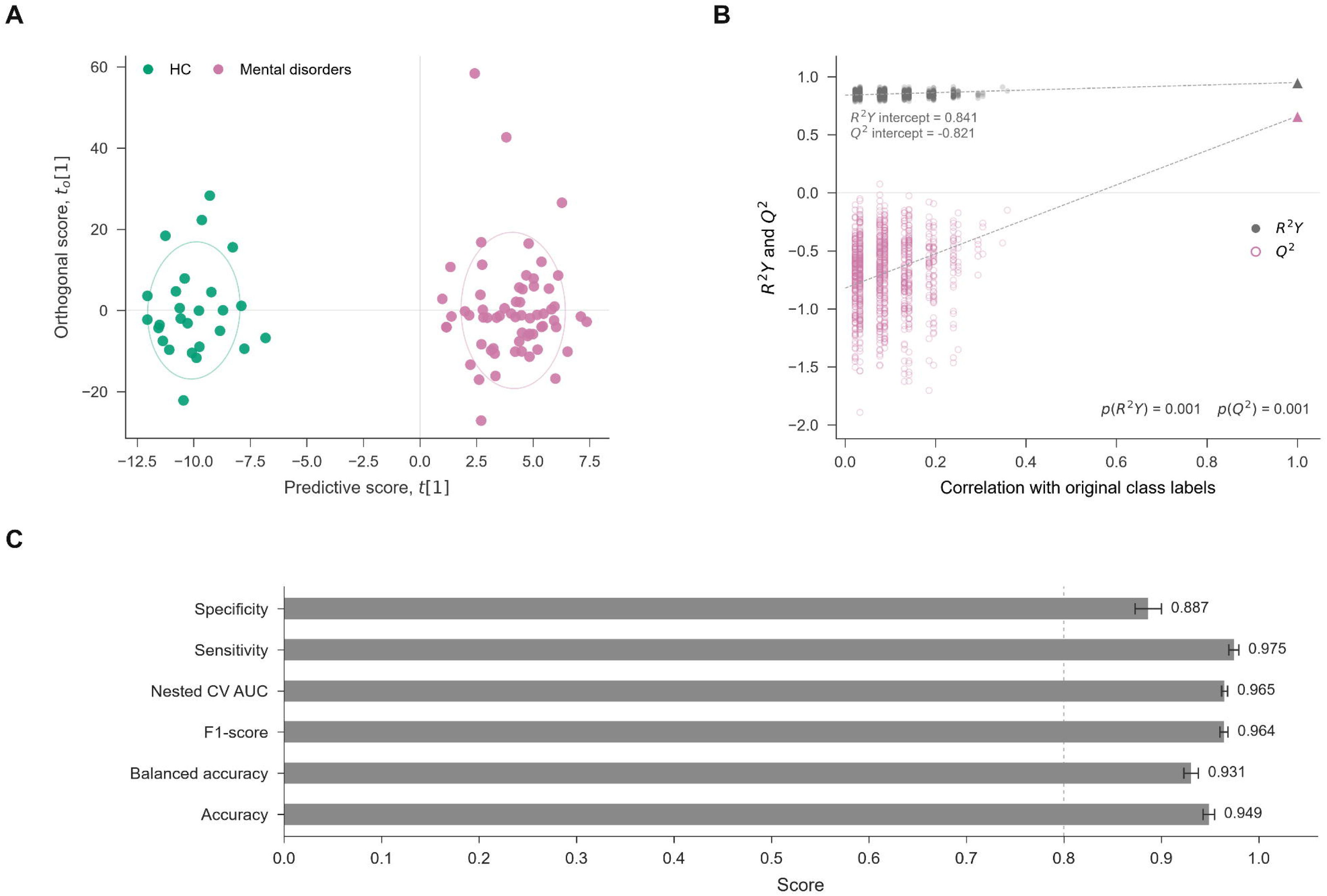
OPLS-DA discrimination, validation, and predictive performance based on serum metabolomic profiles. **(A**) OPLS-DA score plot based on 1,419 serum metabolites showing separation between healthy controls (HC) and participants with mental disorders, comprising major depressive disorder (MDD) and schizophrenia (SCZ). **(B)** Permutation validation of the OPLS-DA model based on 1,000 permutations of the class labels. Permuted R²Y and Q² values are plotted against their correlation with the original class labels, with dashed lines indicating the corresponding regression lines. Triangles at a correlation of 1 represent the original model. Regression intercepts and empirical permutation *P* values are shown within the panel. **(C)** Predictive performance assessed by nested cross-validation, including specificity, sensitivity, area under the receiver operating characteristic curve (AUC), F1-score, balanced accuracy, and accuracy. Bars indicate point estimates, with empirical outer-fold 95% intervals shown where available; the dashed vertical line indicates a reference value of 0.80.

### Selection of the key metabolites and module construction

Feature selection yielded 58 first-stage candidates, of which 38 metabolites were retained after stability selection (ESF, Table S1). The heatmap of the 38 retained metabolites showed group-related patterns across HC, MDD, and SCZ (**Fig. 2B**).

In the locked internal held-out test set, random-forest and elastic-net classifiers based on the 38-metabolite panel achieved AUCs of 0.908 and 0.928, respectively, with an overall accuracy of 0.815 (22/27) for both models (ESF, Fig. S2).

PCA of 20 of the 38 retained metabolites yielded three components, interpreted as PC1 mitochondrial lipid bioenergetics, PC2 sphingolipid stress signalling, and PC3 metabolic resilience (ESF, Table S2). The remaining 18 metabolites formed eight non-PCA biologically informed modules (ESF, Table S3). Together, these three PCA-derived components and eight non-PCA metabolic modules constituted the final set of 11 metabolic modules. Additional PCA diagnostics are provided in ESF, Results.

### Differences in metabolic modules

Significant group differences were observed for all 11 metabolic modules (**Table 2**). Compared with HC, both MDD and SCZ showed lower PC1 mitochondrial lipid bioenergetics and PC3 metabolic resilience and higher PC2 sphingolipid stress signalling, nonate, oxidative lipid damage + LOX, RAAS stress, terpenoid metabolism, and alkamide. Most of these modules did not differ significantly between MDD and SCZ. PUFA membrane remodelling was progressively higher from HC to SCZ to MDD, with significant differences between all three groups, whereas ANTIOXIDANT STATE was lower in MDD than in both HC and SCZ. Taurine was higher in MDD than in HC. The distribution of the 11 metabolic modules across the three diagnostic groups is shown in the heatmap in **Fig. 2C**.

The primary integrated OPLS-DA showed separation between HC and pooled MDD+SCZ (**Fig. 5A**). Eleven predictors had VIP scores >1, with oxidative lipid damage + LOX, PUFA membrane remodelling, and RAAS stress showing the highest VIP scores, followed by emotional abuse and PC2 sphingolipid stress signalling (**Fig. 5B**). Permutation testing yielded a Q² intercept of −0.291, with empirical P = 0.001 for both R²Y and Q² (**Fig. 5C**). Repeated nested cross-validation yielded a specificity of 0.800, sensitivity of 0.929, AUC of 0.946, F1-score of 0.923, balanced accuracy of 0.864, and overall accuracy of 0.891 (**Fig. 5D**). Alternative OPLS-DA configurations varying the inclusion of the two putatively diet-derived modules and ACEs are shown in ESF, Figs. S3–S4. Additional OPLS-DA model characteristics are provided in ESF, Results.

**Figure 5.**
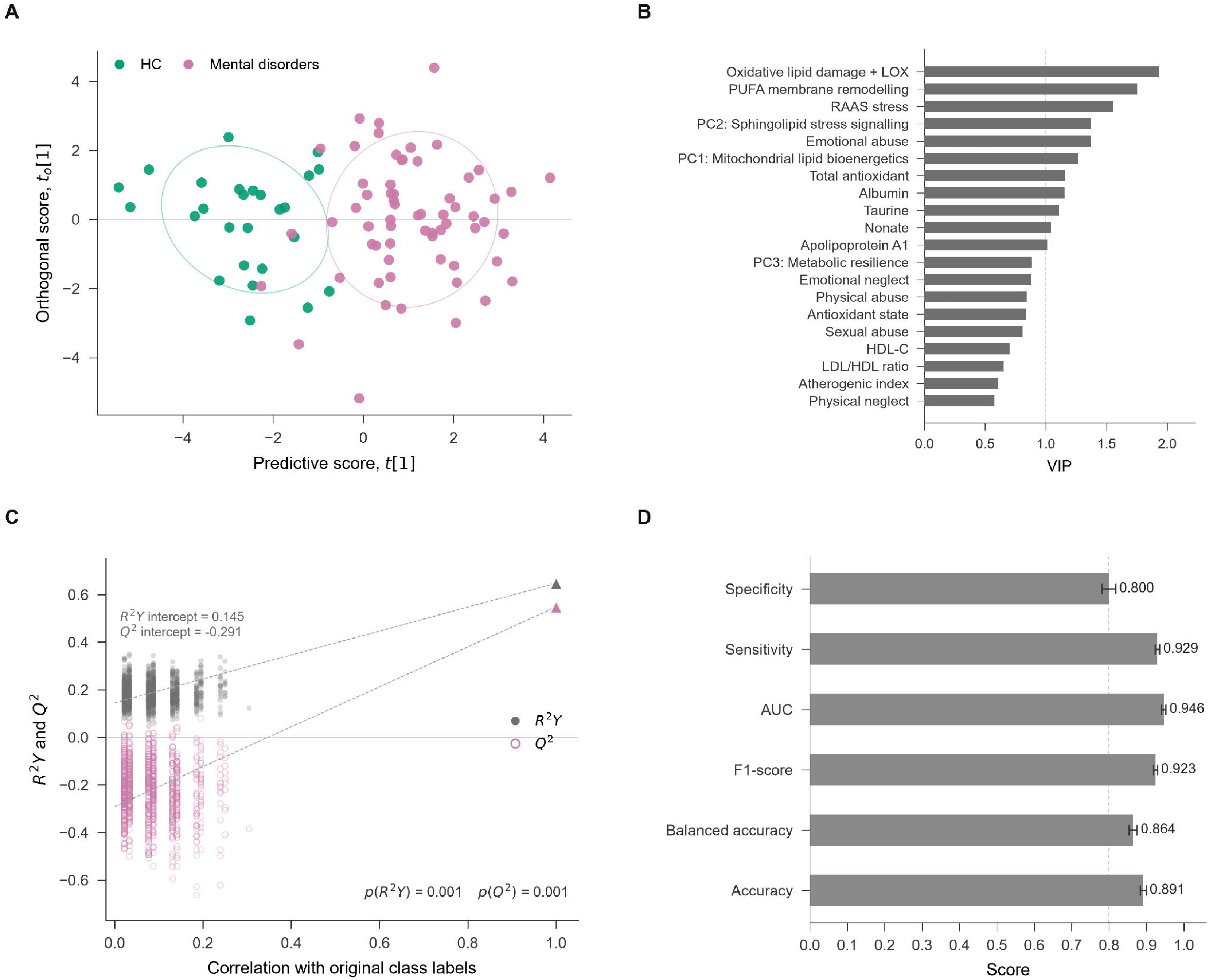
OPLS-DA classification based on metabolic and adverse childhood experience variables. The model included metabolic and adverse childhood experience (ACE) variables while excluding the two plant-derived predictors, terpenoid metabolism and alkamide. **(A)** OPLS-DA score plot showing discrimination between healthy controls (HC) and the pooled major depressive disorder (MDD) and schizophrenia (SCZ) group (labels as mental disorders). The final model contained one predictive and one orthogonal component and used stratified 5-fold cross-validation. Ellipses indicate 1.5-SD group covariance regions for visualisation only. **(B)** Variable importance in projection (VIP) scores for the 20 highest-ranking predictors in the model. Predictors are ordered by decreasing VIP score, and the vertical dashed line indicates VIP = 1.0. **(C)** Permutation validation based on 1,000 permutations of the class labels. Permuted R²Y values are shown as filled gray circles and Q² values as open magenta circles; dashed lines indicate the corresponding regression lines, and triangles at a correlation of 1 represent the original model. Regression intercepts and empirical permutation *P* values are shown within the panel. **(D)** Predictive performance assessed by nested cross-validation. Bars show point estimates for specificity, sensitivity, area under the receiver operating characteristic curve (AUC), F1-score, balanced accuracy, and accuracy; whiskers indicate empirical 95% bootstrap intervals where available, and the vertical dashed line denotes a reference value of 0.80.

### Correlations among ACEs, metabolic and clinical variables

**Figure 6A** shows associations of the metabolic modules with demographic characteristics and childhood adversity. After FDR correction, PC3 metabolic resilience was negatively correlated with emotional abuse, whereas PC2 sphingolipid stress signalling was positively correlated with physical abuse. Oxidative lipid damage + LOX, PUFA membrane remodelling, RAAS stress, and terpenoid metabolism showed positive associations with total childhood-trauma burden and/or abuse domains, with RAAS stress showing the broadest pattern. No FDR-significant associations were observed for PC1 mitochondrial lipid bioenergetics, taurine, nonate, ANTIOXIDANT STATE, or alkamide.

**Figure 6.**
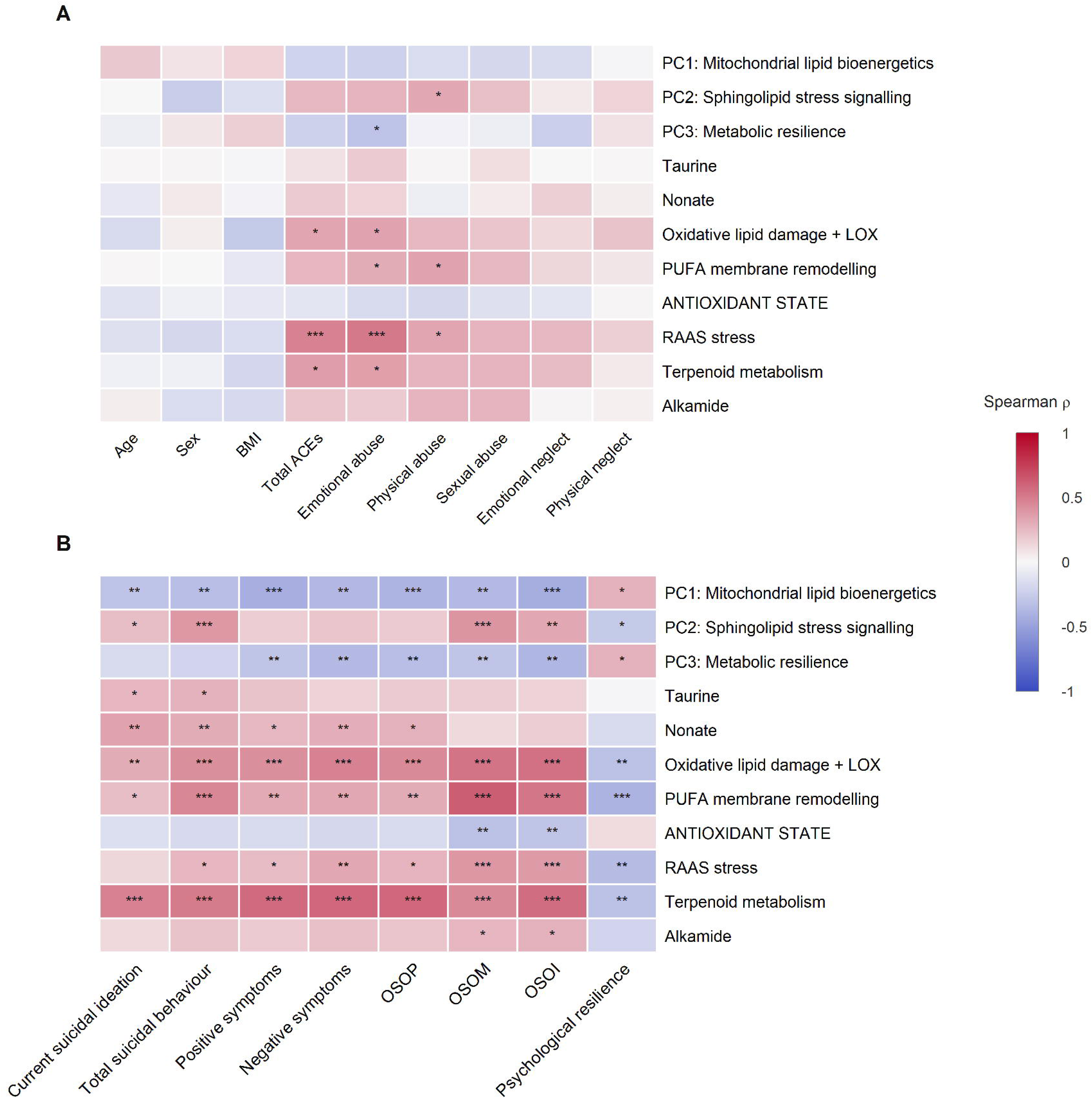
Associations of metabolic modules with demographic characteristics, childhood adversity, and symptom domains. Spearman correlation heatmaps show associations between the 11 metabolic modules and demographic, childhood-adversity, and clinical measures. **(A)** Correlations with age, sex, body mass index (BMI), total adverse childhood experiences (ACEs), and five ACE domains: emotional abuse, physical abuse, sexual abuse, emotional neglect, and physical neglect. **(B)** Correlations with current suicidal ideation, total suicidal behaviour, positive symptoms, negative symptoms, overall severity of psychosis (OSOP), overall severity of mood (OSOM), overall severity of illness (OSOI), and psychological resilience. Cell colour represents Spearman’s ρ, with blue indicating negative correlations, white indicating values near zero, and red indicating positive correlations. P values were adjusted using the Benjamini–Hochberg false-discovery-rate (FDR) procedure separately within each panel. Asterisks denote FDR-adjusted significance: *q* < 0.05 (*), *q* < 0.01 (**), and *q* < 0.001 (***).

Clinical dimensions showed broader metabolic associations (Fig. 6B). PC1 mitochondrial lipid bioenergetics and PC3 metabolic resilience were generally negatively correlated with clinical severity and positively correlated with psychological resilience, whereas PC2 showed largely opposite associations. Oxidative lipid damage + LOX, PUFA membrane remodelling, RAAS stress, and terpenoid metabolism were positively correlated with multiple clinical dimensions and negatively correlated with psychological resilience. Lower ANTIOXIDANT STATE was associated with higher OSOM and OSOI, whereas taurine, nonate, and alkamide showed more selective associations.

### Multiple regression models of clinical outcomes

**Table 3** shows the multiple regression models for the eight clinical outcomes after exclusion of the two putatively diet-derived modules, terpenoid metabolism and alkamide. Current suicidal ideation was positively associated with nonate, PC2 sphingolipid stress signalling, and age, and negatively associated with PC1 mitochondrial lipid bioenergetics (adjusted R² = 0.214). Total suicidal behaviour was positively associated with PC2 and negatively associated with PC1 (adjusted R² = 0.252). Positive symptoms, negative symptoms, and OSOP showed a common pattern of positive associations with oxidative lipid damage + LOX and negative associations with PC1 mitochondrial lipid bioenergetics and PC3 metabolic resilience; total cholesterol was additionally negatively associated with positive symptoms and OSOP. The corresponding adjusted R² values were 0.271, 0.287, and 0.322, respectively.

**Table 3.** Multivariable linear regression models of clinical outcomes excluding putatively plant-derived variables.

| Dependent Variables | Explanatory Variables | Coefficients of input variables |  |  | Model statistics |  |  |  |
| --- | --- | --- | --- | --- | --- | --- | --- | --- |
| | | $\beta$ | $t$ | $P$ | Adj.R<sup>2</sup><br>CV R<sup>2</sup> | $F$<br>df | $P$ | RMSE<br>MAE |
| #1. Current suicidal ideation (z) | **Model** |  |  |  | 0.214 | 6.98 | <0.001 | 0.915 |
|  | Nonate | 0.249 | 2.47 | 0.015 | 0.153 | 4/84 |  | 0.772 |
|  | PC2: Sphingolipid stress signalling | 0.254 | 2.70 | 0.008 |  |  |  |  |
|  | PC1: Mitochondrial lipid bioenergetics | -0.258 | -2.53 | 0.013 |  |  |  |  |
|  | Age | 0.209 | 2.18 | 0.032 |  |  |  |  |
| #2. Total suicidal behaviour (z) | **Model** |  |  |  | 0.252 | 15.81 | <0.001 | 0.882 |
|  | PC1: Mitochondrial lipid bioenergetics | -0.319 | -3.48 | <0.001 | 0.214 | 2/86 |  | 0.717 |
|  | PC2: Sphingolipid stress signalling | 0.405 | 4.42 | <0.001 |  |  |  |  |
| #3. Positive symptoms (z) | **Model** |  |  |  | 0.271 | 9.19 | <0.001 | 0.898 |
|  | PC1: Mitochondrial lipid bioenergetics | -0.328 | -3.43 | <0.001 | 0.220 | 4/84 |  | 0.711 |
| | | $\beta$ | $t$ | $P$ | Adj.R<sup>2</sup><br>CV R<sup>2</sup> | $F$<br>df | $P$ | RMSE<br>MAE |
|  | Oxidative lipid damage + LOX | 0.220 | 2.19 | 0.032 |  |  |  |  |
|  | PC3: Metabolic resilience | -0.242 | -2.47 | 0.016 |  |  |  |  |
|  | Total cholesterol | -0.198 | -2.06 | 0.043 |  |  |  |  |
| #4. Negative symptoms (z) | **Model** |  |  |  | 0.287 | 12.83 | <0.001 | 0.881 |
|  | Oxidative lipid damage + LOX | 0.315 | 3.24 | 0.002 | 0.249 | 3/85 |  | 0.717 |
|  | PC1: Mitochondrial lipid bioenergetics | -0.271 | -2.87 | 0.005 |  |  |  |  |
|  | PC3: Metabolic resilience | -0.262 | -2.78 | 0.007 |  |  |  |  |
| #5. OSOP (z) | **Model** |  |  |  | 0.322 | 11.45 | <0.001 | 0.865 |
|  | Oxidative lipid damage + LOX | 0.262 | 2.70 | 0.008 | 0.275 | 4/84 |  | 0.701 |
|  | PC1: Mitochondrial lipid bioenergetics | -0.315 | -3.42 | <0.001 |  |  |  |  |
| | | $\beta$ | $t$ | $P$ | Adj.R<sup>2</sup><br>CV R<sup>2</sup> | $F$<br>df | $P$ | RMSE<br>MAE |
|  | PC3: Metabolic resilience | -0.288 | -3.04 | 0.003 |  |  |  |  |
|  | Total cholesterol | -0.196 | -2.11 | 0.038 |  |  |  |  |
| #6. OSOM (z) | **Model** |  |  |  | 0.525 | 49.57 | <0.001 | 0.700 |
|  | PUFA membrane remodelling | 0.512 | 6.84 | <0.001 | 0.505 | 2/86 |  | 0.586 |
|  | Emotional abuse | 0.415 | 5.54 | <0.001 |  |  |  |  |
| #7. OSOI (z) | **Model** |  |  |  | 0.606 | 23.54 | <0.001 | 0.668 |
|  | Oxidative lipid damage + LOX | 0.234 | 2.43 | 0.017 | 0.560 | 6/82 |  | 0.514 |
|  | PC1: Mitochondrial lipid bioenergetics | -0.306 | -4.32 | <0.001 |  |  |  |  |
|  | PC3: Metabolic resilience | -0.203 | -2.84 | 0.006 |  |  |  |  |
|  | Sexual abuse | 0.207 | 2.95 | 0.004 |  |  |  |  |
| | | $\beta$ | $t$ | $P$ | Adj.R<sup>2</sup><br>CV R<sup>2</sup> | $F$<br>df | $P$ | RMSE<br>MAE |
|  | PUFA membrane remodelling | 0.253 | 2.70 | 0.008 |  |  |  |  |
|  | Emotional neglect | 0.190 | 2.66 | 0.009 |  |  |  |  |
| #8. Psychological resilience (z) | **Model** |  |  |  | 0.386 | 12.05 | <0.001 | 0.825 |
|  | Emotional abuse | -0.336 | -3.70 | <0.001 | 0.324 | 5/83 |  | 0.667 |
|  | PUFA membrane remodelling | -0.378 | -4.10 | <0.001 |  |  |  |  |
|  | Age | 0.238 | 2.68 | 0.009 |  |  |  |  |
|  | Log free cholesterol | -0.257 | -2.99 | 0.004 |  |  |  |  |
|  | Taurine | 0.195 | 2.13 | 0.037 |  |  |  |  |

$\beta$ denotes the standardised regression coefficient. Predictive performance was assessed using 10-fold cross-validation repeated 200 times with the selected predictor set held fixed. CV R<sup>2</sup>, RMSE, and MAE were calculated from aggregated out-of-fold predictions. Coefficient stability was assessed using 1,000 participant-level bootstrap resamples; all retained coefficients had 95% bootstrap intervals excluding zero.
Multicollinearity was low in all final models (maximum VIF=2.05; conservative criterion VIF<2.5).
Abbreviations: CTQ-SF, Childhood Trauma Questionnaire-Short Form; CV, cross-validation; LOX, lipoxigenase; MAE, mean absolute error; OSOI, overall severity of illness; OSOM, overall severity of mood; OSOP,
overall severity of psychosis; PC, principal component; PUFA, polyunsaturated fatty acid; RAAS, renin-angiotensin-aldosterone system; RMSE, root-mean-square error; VIF, variance inflation factor; z, standardised score.

OSOM was positively associated with PUFA membrane remodelling and emotional abuse (adjusted R² = 0.525). OSOI was positively associated with oxidative lipid damage + LOX, sexual abuse, PUFA membrane remodelling, and emotional neglect, and negatively associated with PC1 mitochondrial lipid bioenergetics and PC3 metabolic resilience (adjusted R² = 0.606). Psychological resilience was negatively associated with emotional abuse, PUFA membrane remodelling, and log-transformed free cholesterol, and positively associated with age and taurine (adjusted R² = 0.386). All predictors retained in the primary regression models showed bootstrap stability, and multicollinearity was low (maximum VIF = 2.05).

In exploratory models allowing the two putatively diet-derived modules to enter, terpenoid metabolism was retained in six of the eight clinical-outcome models and was the sole retained predictor of current suicidal ideation, positive symptoms, negative symptoms, and OSOP. Neither terpenoid metabolism nor alkamide was retained in the OSOM or psychological-resilience models.

### Effects of putative confounding variables

After adjustment for age, sex, and BMI, significant diagnostic-group effects were retained for all eleven metabolic modules, with the principal group-difference patterns preserved. In medication sensitivity analyses restricted to participants with MDD or SCZ, current antidepressant, benzodiazepine, mood-stabiliser, and antipsychotic use showed no significant multivariate associations with the metabolic or biochemical variables, and no univariate association remained significant after Benjamini–Hochberg correction.

## Discussion

### Transdiagnostic Metabolic Architecture in MDD and SCZ

The first major finding was a broadly similar pattern of serum metabolic differences in MDD and SCZ relative to HC. Relative to controls, both mental disorders were characterised by reduced mitochondrial lipid bioenergetics and metabolic resilience, together with increased sphingolipid stress signalling, oxidative lipid damage, and LOX-related metabolism, and RAAS stress. This broadly similar pattern was accompanied by selective diagnostic divergence: PUFA membrane remodelling increased progressively from controls to SCZ to MDD, whereas ANTIOXIDANT STATE was selectively reduced in MDD.

Previous metabolomic and lipidomic studies likewise indicate substantial molecular overlap between MDD and SCZ with a smaller set of disorder-dependent features [5–7]. Our findings extend this literature by showing, within the same serum framework, a shared metabolic core with superimposed diagnostic divergence across bioenergetic, lipid, oxidative, and stress-related domains. Prior studies have documented lipid peroxidation and altered antioxidant defences in both depression and psychosis, with substantial heterogeneity across individual antioxidant systems [13, 37, 38].

Childhood adversity also intersected with this transdiagnostic metabolic architecture. Emotional abuse showed the broadest pattern of associations, whereas physical abuse and total childhood-trauma score showed more selective relationships. This extends prior evidence linking childhood trauma to psychiatric outcomes and NIMETOX pathways [2, 15–17]. RAAS may form one component of a biological embedding process, as childhood trauma, particularly abuse, has been associated with higher circulating aldosterone [20], paralleling the ACE-related RAAS associations observed here. These data suggest a psychosocial–metabolic association between early adversity and distributed stress-related biology.

### PUFA remodelling and emotional abuse across mood and resilience

The second major finding is the recurrent involvement of PUFA membrane remodelling and emotional abuse across mood severity and psychological resilience. Greater OSOM was independently associated with greater PUFA membrane remodelling and emotional abuse, whereas both variables were inversely associated with psychological resilience. This recurring pattern suggests a cross-dimensional psychosocial–metabolic configuration spanning greater mood disturbance and lower psychological resilience.

MDD has frequently been associated with altered circulating PUFA composition and broader abnormalities across phospholipid and membrane lipid classes [10, 11, 21].

Importantly, this PUFA remodelling composite should not be equated with deficiency or excess of any individual omega-3 or omega-6 fatty acid. It is more appropriately interpreted as a metabolic correlate of PUFA-related membrane remodelling, with potential relevance to membrane organisation, lipid-mediator biosynthesis, and cell signalling [9].

Emotional abuse formed the complementary psychosocial component. Meta-analytic and transdiagnostic evidence links childhood maltreatment—particularly emotional abuse—to MDD, affective instability, and emotion dysregulation [39, 40]. Human metabolomic studies also suggest that childhood adversity can be accompanied by alterations in fatty-acid and other metabolic pathways [18, 19, 35]. Longitudinal evidence further indicates that childhood trauma and lower omega-3 composition independently predicted persistence of depressive symptoms despite being unrelated to one another [41].

### Bioenergetic–Redox–Sphingolipid Signatures of Psychosis and Suicidality

The third major finding is that positive and negative symptoms, and OSOP were consistently associated with greater oxidative lipid damage and LOX-related metabolism together with lower mitochondrial lipid bioenergetics and metabolic resilience. By contrast, both current suicidal ideation and total suicidal behaviour were associated with lower mitochondrial lipid bioenergetics and higher sphingolipid stress signalling. Lower mitochondrial lipid bioenergetics therefore represented the principal shared feature; psychosis-related models additionally included greater redox stress and lower metabolic resilience, whereas suicidal models additionally included higher sphingolipid stress signalling.

Systematic studies have reported disturbances in oxidative phosphorylation and bioenergetic regulation in schizophrenia [42]. Meta-analytic and first-episode studies also indicate increased lipid peroxidation, with some oxidative indices related to positive symptom severity [3, 4, 14, 22]. Suicidality showed a related but distinct bioenergetic–sphingolipid configuration. Metabolomic studies of suicidal depression have implicated mitochondrial and lipid-related pathways, including alterations in ceramide and sphingomyelin species [23], while transcriptomic work has identified suicide-associated changes in mitochondrial genes [43]. Our PC2 sphingolipid stress signalling module is best considered a composite correlate of sphingolipid-related stress rather than evidence of increased ceramide per se.

### Putatively diet-derived biotransformation signals and exposome sensitivity

The fourth major finding is that terpenoid metabolism was retained in six of the eight exploratory multivariable models, including current suicidal ideation, total suicidal behaviour, positive symptoms, negative symptoms, OSOP, and OSOI. If entered in the analysis, it was the sole retained predictor of current suicidal ideation, positive symptoms, negative symptoms, and OSOP. Neither terpenoid metabolism nor alkamide was retained in the OSOM or psychological-resilience models.

Circulating terpenoid-related compounds may reflect dietary or environmental exposures [25] as well as gut microbial and hepatic biotransformation [44, 45]. In China, p-menthan-2-one (carvomenthone; FEMA 3176) is an approved food flavouring substance and exposure can occur through flavoured processed foods, confectionery, beverages, mint-flavoured products, and flavour preparations. In our Chinese patients, these results may suggest alterations in food/flavouring exposure, intestinal absorption, microbial transformation, and hepatic xenobiotic metabolism yielding a differential serum terpenoid profile. Nevertheless, the terpenoid-related finding should be considered biologically exploratory and requires direct exposure characterisation and independent replication.

### Metabolic and psychosocial architecture of psychological resilience

The fifth major finding is that psychological resilience was associated with a partially distinct multidimensional psychosocial–metabolic architecture. The resilience model showed that greater PUFA membrane remodelling, emotional abuse, and higher free cholesterol were associated with lower psychological resilience, whereas older age and higher taurine were associated with greater resilience. Radford-Smith et al. identified a multivariate blood metabolite profile associated with susceptibility and resilience to MDD, with pyruvate and lactate among the prominent contributors [24]. Physiological evidence links taurine to mitochondrial function, calcium and osmotic regulation, and redox homeostasis [46], while peripheral metabolomic and preclinical studies support its relevance to depression-related phenotypes [47, 48]. Cerebral cholesterol homeostasis is largely regulated independently of peripheral cholesterol, although peripheral cholesterol metabolism may influence brain function through immune-inflammatory and vascular pathways, oxysterol signalling, and blood–brain barrier dysfunction [1, 49].

### Limitations

This cross-sectional study does not permit causal inference or distinction between state- and trait-related metabolic effects. The results deserve replication in independent cohorts from other cultures and countries. Future longitudinal and externally validated studies should examine these metabolic architectures alongside additional NIMETOX-related pathways, and environmental exposures. Residual confounding by diet, lifestyle, and other environmental exposures is possible. Metabolite identities based on untargeted annotation remain putative unless confirmed with authentic standards. Serum metabolic alterations cannot be assumed to directly reflect corresponding processes in the brain, and the biological origin of the diet-derived signals remains uncertain.

## Conclusion

This study identified broadly similar serum metabolic patterns in MDD and SCZ relative to controls. Key features included mitochondrial lipid bioenergetics, metabolic resilience, sphingolipid stress signalling, oxidative lipid damage, and LOX-related metabolism, PUFA membrane remodelling, and RAAS stress. PUFA remodelling and emotional abuse converged across mood severity, whereas psychotic and suicidal dimensions showed partially overlapping bioenergetic–redox–lipid configurations.

Psychological resilience was associated with a partially distinct psychosocial–metabolic architecture. These multivariable patterns suggest that clinically informative signatures may involve combinations of metabolic, biochemical, and psychosocial features rather than single biomarkers. Future prospective studies should determine whether such multidomain profiles can identify clinically meaningful metabolic phenotypes and whether they are associated with treatment response, illness course, or prognosis.

## Data Availability

All data produced in the present study are available upon reasonable request to the authors

## Ethics approval and consent to participate

This study was approved by the Ethics Committee of the University of Electronic Science and Technology of China (approval no. 30850) and conducted in accordance with the Declaration of Helsinki. Written informed consent was obtained from all participants.

## Author contributions

Yueyang Luo: Methodology, Visualization, Writing – original draft. Hongzhou Wu: Investigation, Methodology. Chenghui Yang: Investigation. Tangcong Chen: Methodology. Abbas F. Almulla: Methodology. Yingqian Zhang: Conceptualization, Visualization, Writing – review & editing. Michael Maes: Conceptualization, Formal analysis, Supervision, Writing – original draft, Writing – review & editing.

## Data availability

The dataset supporting this study is available from the corresponding author (MM) upon reasonable request and after a thorough data review.

## Acknowledgements

Not applicable.

