## Supplementary Material for "Transdiagnostic metabolic architecture across major depressive disorder and schizophrenia: a NIMETOX systems-biology approach"

**ELECTRONIC SUPPLEMENTARY FILE (ESF)**

**ESF, Methods.**

**Method S1. Participants and clinical phenotypes**

The final metabolomics cohort comprised 89 participants: 26 healthy controls (HC), 34 with major depressive disorder (MDD), and 29 with schizophrenia (SCZ), aged 18–65 years. MDD and SCZ were diagnosed by a senior psychiatrist according to DSM-5 criteria using a structured diagnostic interview; the Mini-International Neuropsychiatric Interview (M.I.N.I. 6.0) was additionally administered to assess psychiatric diagnoses and comorbidity (American Psychiatric Association, 2013; Sheehan et al., 1998). Exclusion criteria included other major psychiatric disorders, borderline and antisocial personality disorder or intellectual disability, neurological or major medical illness, pregnancy or breastfeeding, recent severe allergy or infection, current immunosuppressive or immunomodulatory treatment, therapeutic antioxidant or omega-3 supplementation within the preceding 3 months, recent surgery, frequent analgesic use, and traumatic brain injury with loss of consciousness. Current psychotropic medication use was recorded at assessment.

Suicidality was assessed using the Columbia-Suicide Severity Rating Scale (C-SSRS) (Posner et al., 2011), positive and negative symptoms using the Positive and Negative Syndrome Scale (PANSS) (Kay et al., 1987), depressive symptoms using the Beck Depression Inventory-II (BDI-II) (Beck et al., 2011), and state anxiety using the State-Trait Anxiety Inventory, state version (STAI-state) (Skapinakis, 2014). Current suicidal ideation was represented by the first principal component extracted from six C-SSRS items assessing current ideation severity and intensity. Lifetime suicidal behaviour was calculated as a standardised composite of lifetime suicidal ideation and suicide attempts, and total suicidal behaviour combined standardised lifetime suicidal behaviour with standardised current suicidal ideation (Maes et al., 2024). Overall severity of mood (OSOM) was represented by the first principal component extracted from BDI-II, STAI-state, and binary MDD diagnostic status. Overall severity of psychosis (OSOP) was calculated as z (PANSS positive) + z (PANSS negative), and overall severity of illness (OSOI) as [zOSOM + zOSOP]. Psychological resilience was assessed using the Connor–Davidson Resilience Scale (CD-RISC) (Connor & Davidson, 2003). Items 2, 3, 9, and 20 were excluded because they did not meet the loading criterion, and resilience was represented by the first principal component extracted from the remaining 21 items. Adverse childhood experiences (ACEs) were assessed using the Childhood Trauma Questionnaire–Short Form (CTQ-SF) (Bernstein et al., 2003). Five domains were analysed: emotional abuse, physical abuse, sexual abuse, emotional neglect, and physical neglect. Total childhood trauma was calculated as the sum of the five domain scores. Standardised domain scores were used in downstream analyses as specified.

**Method S2. Biochemical measures**

Fasting venous blood (10 mL) was collected between 07:00 and 08:00 h, centrifuged at 3500 rpm for 10 min, and serum aliquots were stored at −80 °C until analysis. Serum albumin, triglycerides (TG), total cholesterol (TC), LDL-C, HDL-C, apolipoprotein A1 (ApoA1), and free cholesterol (FC) were measured using established assays on an ADVIA 2400 automated biochemical analyser (Siemens Healthcare Diagnostics). FC was log10-transformed before subsequent analyses. The LDL-C/HDL-C ratio was calculated directly from the corresponding concentrations. The atherogenic index of plasma (AIP) followed the previously applied z-unit formulation in this cohort: AIP = z(TG) − z(HDL-C) (Maes et al., 2024). The antioxidant-defence composite was calculated as z[z(albumin) + z(HDL-C) + z(ApoA1)], with higher values indicating a more favourable combined antioxidant-related profile. This biochemical composite was distinct from ANTIOXIDANT STATE, which additionally incorporated three selected metabolite features.

**Method S3. Metabolomics**

Untargeted serum metabolomics was performed using an established workflow with study-specific modifications. Serum samples were extracted with acidified methanol, centrifuged, and the resulting supernatants were used for LC–MS analysis. Extraction solvent served as a blank, and pooled quality-control (QC) samples prepared from study samples were injected regularly throughout the analytical sequence to monitor analytical stability. Metabolomic profiling was performed using an ACQUITY UPLC system (Waters) coupled to a Q-Exactive Plus high-resolution mass spectrometer (Thermo Fisher Scientific). Data were acquired in separate positive- and negative-electrospray-ionisation modes using full MS/data-dependent MS² acquisition over m/z 80–1200. Raw data were converted to mzML format and processed in Progenesis QI version 2.4 for peak detection, alignment, retention-time correction, missing-value handling, and signal correction. Metabolites were annotated against an in-house spectral library, the Human Metabolome Database (HMDB), and the Kyoto Encyclopedia of Genes and Genomes (KEGG). Features with an annotation score >0.5 and QC coefficient of variation <0.30 were retained. Positive- and negative-ion datasets were merged after resolving duplicate annotations, yielding 1,419 annotated metabolite features for subsequent analyses.

**Method S4. Metabolite screening and feature selection**

The 1,419 annotated metabolite features were subjected to a study-specific, rule-based screen of HMDB annotations to generate an endogenous-enriched candidate pool. Features annotated as microbial, drug-related, toxic, or pollutant-associated were excluded, whereas endogenous features were retained. Features with mixed endogenous and food- or plant-associated annotations were retained only when their HMDB descriptions supported an endogenous biochemical role. This procedure reduced the dataset to 540 candidate metabolites. Candidate abundances were centred-log-ratio (CLR) transformed for inferential and multivariate feature-selection analyses, whereas untransformed abundances were retained for fold-change estimation.

Supervised feature selection for discrimination of HC from pooled MDD+SCZ was restricted to a diagnosis-stratified 70% feature-selection subset (n=62). The remaining 30% of participants (n=27) were not used during supervised feature selection. Four complementary approaches were applied: Wilcoxon rank-sum testing, partial least-squares discriminant analysis (PLS-DA), random forest, and LASSO logistic regression. The union of features selected by these procedures yielded 58 first-stage candidates. The 58 candidates were subsequently evaluated using repeated PLS-DA and random-forest resampling within the same feature-selection subset. Final retention required concordant stability across both procedures, with a selection frequency ≥0.50 in each. This procedure yielded 38 retained metabolites. Following feature selection, the retained metabolites were returned to the full cohort (n=89) for construction of the biologically informed metabolic modules and downstream analyses.

**Method S5. Construction of functional metabolic modules**

The 38 retained metabolites were returned to the full cohort for construction of the final metabolic modules. Values below the 5th percentile or above the 95th percentile were winsorized to the corresponding percentile, and metabolite abundances were standardised as required for PCA and composite-score construction. Twenty metabolites were entered into principal component analysis (PCA) with Varimax rotation and Kaiser normalization. Metabolites were assigned to the component with the largest absolute loading, and retained components were interpreted according to their dominant loadings and biological composition.

The remaining 18 metabolites were summarised into eight biologically informed modules: oxidative lipid damage + LOX, RAAS stress, PUFA membrane remodelling, ANTIOXIDANT STATE, taurine, nonate, terpenoid metabolism, and alkamide. ANTIOXIDANT STATE combined three selected metabolite features with ApoA1, HDL-C, and albumin. Terpenoid metabolism and alkamide were classified as putatively diet-derived, exposure-sensitive modules based on HMDB annotations; this classification did not imply direct measurement of dietary or environmental exposure.

**Method S6. OPLS-DA and model validation**

OPLS-DA was used to discriminate HC from pooled MDD+SCZ using either all 1,419 annotated metabolite features or an integrated predictor set. The primary integrated model comprised nine non-diet-derived metabolic modules, 12 biochemical measures (albumin, transferrin, TG, TC, HDL-C, LDL-C, VLDL-C, ApoA1, log-transformed FC, LDL-C/HDL-C ratio, antioxidant-defence composite, and AIP), and the five CTQ-SF childhood-adversity domains; terpenoid metabolism and alkamide were excluded. Model significance was assessed using 1,000 label permutations, and predictive performance using repeated nested five-fold cross-validation. VIP scores were obtained from the full-data model, and score-plot ellipses were used for visualisation only.

Three alternative integrated configurations varied the inclusion of the two putatively diet-derived modules and the childhood-adversity domains. The 38-metabolite panel was additionally evaluated using random-forest and elastic-net classifiers trained in the feature-selection subset (n=62) and tested in the locked held-out subset (n=27).

**Method S7. Correlation and regression analyses**

Spearman rank correlations were used to examine associations of the 11 biologically informed metabolic modules with age, sex, rank-transformed BMI, total childhood-trauma score, the five CTQ-SF domains, and eight clinical measures: current suicidal ideation, total suicidal behaviour, positive symptoms, negative symptoms, OSOP, OSOM, OSOI, and psychological resilience. P values were adjusted using the Benjamini–Hochberg false-discovery-rate procedure separately within the demographic/childhood-adversity and clinical panels.

Stepwise linear regression was used to examine multivariable associations with the eight clinical outcomes. Candidate predictors comprised the biologically informed metabolic modules, the five CTQ-SF domains, biochemical variables, age, sex, and rank-transformed BMI. The primary models excluded terpenoid metabolism and alkamide, whereas exploratory models allowed these variables to enter the selection procedure. Age was excluded a priori from the OSOM candidate set, and sex from the psychological-resilience candidate set. Missing predictor values were replaced by the corresponding median. Model performance and coefficient stability were assessed using repeated 10-fold cross-validation and 1,000 participant-level bootstrap resamples, and multicollinearity was assessed using variance inflation factors.

**ESF, Results.**

The first principal component used to derive OSOM explained 82.81% of the variance (Kaiser–Meyer–Olkin statistic = 0.636), with all loadings >0.873.

Sampling adequacy for principal component analysis of the 20 selected metabolites was supported by a Kaiser–Meyer–Olkin statistic of 0.821 and a significant Bartlett’s test of sphericity (χ² = 2020.344, df = 190, P < 0.001). Three Varimax-rotated components were retained, together explaining 70.079% of the variance (PC1, 39.559%; PC2, 17.688%; PC3, 12.831%). Based on their dominant loadings and biological composition, these components were interpreted as PC1 mitochondrial lipid bioenergetics, PC2 sphingolipid stress signalling, and PC3 metabolic resilience. Together with the eight non-PCA modules, these components constituted the final set of 11 biologically informed metabolic modules.

The full-metabolome OPLS-DA contained one predictive and three orthogonal components, whereas the primary integrated OPLS-DA contained one predictive and one orthogonal component.

**ESF, Table S1. The 38 metabolites retained after screening and their allocation to subsequent analyses.**

| **HMDB ID** | **Metabolite** | **Analytical allocation** | **Derived variable** | **Scoring approach** |
| --- | --- | --- | --- | --- |
| HMDB0000251 | Taurine | Individual metabolite | Taurine | Standardised abundance |
| HMDB0000909 | trans-4-Hydroxycyclohexylacetic acid | PCA-associated | PC3: Metabolic resilience | PCA-derived score |
| HMDB0000991 | DL-2-Aminooctanoic acid | PCA-associated | PC3: Metabolic resilience | PCA-derived score |
| HMDB0006294 | 16-Hydroxyhexadecanoic acid | PCA-associated | PC2: Sphingolipid stress signalling | PCA-derived score |
| HMDB0010357 | Tetrahydroaldosterone-3-glucuronide | Derived metabolic variable | RAAS stress | Negative contribution |
| HMDB0010404 | LysoPC(22:6(4Z,7Z,10Z,13Z,16Z,19Z)/0:0) | Derived metabolic variable | PUFA membrane remodelling | Positive contribution |
| HMDB0010722 | (R)-3-Hydroxyoctanoic acid | PCA-associated | PC3: Metabolic resilience | PCA-derived score |
| HMDB0010730 | 3-Oxotetradecanoic acid | PCA-associated | PC1: Mitochondrial lipid bioenergetics | PCA-derived score |
| HMDB0011717 | Nonate | Individual metabolite | Nonate | Standardised abundance |
| HMDB0011759 | Cer(d18:0/14:0) | PCA-associated | PC2: Sphingolipid stress signalling | PCA-derived score |
| HMDB0030951 | (E,E)-2,4-Decadienoic isobutylamide | Putatively diet-derived | Alkamide | Standardised abundance |
| HMDB0032369 | L-Menthyl acetoacetate | Putatively diet-derived | Terpenoid metabolism | Negative contribution |
| HMDB0035272 | p-Menthan-2-one | Putatively diet-derived | Terpenoid metabolism | Positive contribution |
| HMDB0035337 | Sterebin A | Putatively diet-derived | Terpenoid metabolism | Negative contribution |
| HMDB0039893 | (1R,2R,4S)-p-Menthane-1,2,8-triol | Putatively diet-derived | Terpenoid metabolism | Negative contribution |
| HMDB0041600 | Ethyl (±)-3-hydroxyoctanoate | PCA-associated | PC1: Mitochondrial lipid bioenergetics | PCA-derived score |
| HMDB0060101 | 12-Hydroxyarachidonic acid | Derived metabolic variable | Oxidative lipid damage + LOX | Positive contribution |
| HMDB0061643 | 3-Carboxy-4-methyl-5-pentyl-2-furanpropanoic acid (CMPF) | Derived metabolic variable | ANTIOXIDANT STATE | Positive contribution |
| HMDB0061656 | 3-Hydroxytetradecanoic acid | PCA-associated | PC1: Mitochondrial lipid bioenergetics | PCA-derived score |
| HMDB0240603 | LysoPS(18:1(9Z)/0:0) | PCA-associated | PC2: Sphingolipid stress signalling | PCA-derived score |
| HMDB0240791 | 3-Methylnonanoylcarnitine | PCA-associated | PC1: Mitochondrial lipid bioenergetics | PCA-derived score |
| HMDB0241044 | 5-Methylheptanoylcarnitine | PCA-associated | PC1: Mitochondrial lipid bioenergetics | PCA-derived score |
| HMDB0241063 | 9-Hydroxydecanoylcarnitine | PCA-associated | PC1: Mitochondrial lipid bioenergetics | PCA-derived score |
| HMDB0241306 | 9-Hydroxydodecanoylcarnitine | PCA-associated | PC1: Mitochondrial lipid bioenergetics | PCA-derived score |
| HMDB0241308 | Tridecanoylcarnitine | PCA-associated | PC1: Mitochondrial lipid bioenergetics | PCA-derived score |
| HMDB0241362 | (12E)-10-Hydroxytetradec-12-enoylcarnitine | PCA-associated | PC1: Mitochondrial lipid bioenergetics | PCA-derived score |
| HMDB0241694 | 5-Hydroxyoctanoylcarnitine | PCA-associated | PC1: Mitochondrial lipid bioenergetics | PCA-derived score |
| HMDB0242462 | (2-Amino-3-hydroxyoctadecyl) dihydrogen phosphate | PCA-associated | PC2: Sphingolipid stress signalling | PCA-derived score |
| HMDB0244521 | 13,16-Docosadienoic acid | Derived metabolic variable | PUFA membrane remodelling | Positive contribution |
| HMDB0245425 | 2,3,4-Trihydroxybutanoic acid | Derived metabolic variable | ANTIOXIDANT STATE | Positive contribution |
| HMDB0247790 | C18-Sphingosine 1-phosphate; D-erythro-Sphingosine-1-phosphate | PCA-associated | PC2: Sphingolipid stress signalling | PCA-derived score |
| HMDB0250928 | Decylubiquinone | PCA-associated | PC1: Mitochondrial lipid bioenergetics | PCA-derived score |
| HMDB0255357 | 2-(20-Hydroxyicosa-5,14-dienoylamino)acetic acid | Derived metabolic variable | ANTIOXIDANT STATE | Positive contribution |
| HMDB0258895 | Tetrahydroaldosterone | Derived metabolic variable | RAAS stress | Positive contribution |
| HMDB0260291 | 5-(4-Carboxybutylperoxy)pentanoic acid | Derived metabolic variable | Oxidative lipid damage + LOX | Positive contribution |
| HMDB0260560 | MG(0:0/18:3(9,11,15)-OH(13)/0:0) | PCA-associated | PC1: Mitochondrial lipid bioenergetics | PCA-derived score |
| HMDB0265376 | PA(PGJ2/20:5(5Z,8Z,11Z,14Z,17Z)) | Derived metabolic variable | PUFA membrane remodelling | Positive contribution |
| HMDB0267690 | PA(20:5(6E,8Z,11Z,14Z,17Z)-OH(5)/i-15:0) | Derived metabolic variable | PUFA membrane remodelling | Positive contribution |

The 38 metabolites retained after screening were allocated to PCA-associated variables, derived metabolic variables, individual metabolites, or putatively diet-derived variables for subsequent analyses. Positive and negative contributions indicate the direction in which standardised metabolite abundances entered the corresponding composite score.

**Abbreviations:** HMDB, Human Metabolome Database; LOX, lipoxygenase; PC, principal component; PCA, principal component analysis; PUFA, polyunsaturated fatty acid; RAAS, renin–angiotensin–aldosterone system.

**ESF, Table S2. Varimax-rotated component loadings of the 20 metabolites included in principal component analysis.**

| **HMDB ID** | **Metabolite** | **PC1 loading** | **PC2 loading** | **PC3 loading** | **Principal assignment** | **Communality** |
| --- | --- | --- | --- | --- | --- | --- |
| HMDB0000909 | trans-4-Hydroxycyclohexylacetic acid | 0.068 | -0.070 | **0.928** | PC3: Metabolic resilience | 0.871 |
| HMDB0000991 | DL-2-Aminooctanoic acid | 0.129 | -0.111 | **0.825** | PC3: Metabolic resilience | 0.710 |
| HMDB0006294 | 16-Hydroxyhexadecanoic acid | -0.092 | **0.770** | -0.138 | PC2: Sphingolipid stress signalling | 0.620 |
| HMDB0010722 | (R)-3-Hydroxyoctanoic acid | 0.052 | 0.005 | **0.876** | PC3: Metabolic resilience | 0.770 |
| HMDB0010730 | 3-Oxotetradecanoic acid | **0.836** | -0.013 | -0.026 | PC1: Mitochondrial lipid bioenergetics | 0.700 |
| HMDB0011759 | Cer(d18:0/14:0) | -0.052 | **0.730** | 0.147 | PC2: Sphingolipid stress signalling | 0.557 |
| HMDB0041600 | Ethyl (±)-3-hydroxyoctanoate | **0.723** | -0.034 | 0.069 | PC1: Mitochondrial lipid bioenergetics | 0.529 |
| HMDB0061656 | 3-Hydroxytetradecanoic acid | **0.731** | 0.140 | -0.099 | PC1: Mitochondrial lipid bioenergetics | 0.564 |
| HMDB0240603 | LysoPS(18:1(9Z)/0:0) | -0.137 | **0.742** | -0.065 | PC2: Sphingolipid stress signalling | 0.574 |
| HMDB0240791 | 3-Methylnonanoylcarnitine | **0.895** | -0.215 | 0.015 | PC1: Mitochondrial lipid bioenergetics | 0.847 |
| HMDB0241044 | 5-Methylheptanoylcarnitine | **0.880** | -0.251 | -0.008 | PC1: Mitochondrial lipid bioenergetics | 0.837 |
| HMDB0241063 | 9-Hydroxydecanoylcarnitine | **0.804** | -0.192 | 0.088 | PC1: Mitochondrial lipid bioenergetics | 0.691 |
| HMDB0241306 | 9-Hydroxydodecanoylcarnitine | **0.940** | -0.061 | 0.054 | PC1: Mitochondrial lipid bioenergetics | 0.890 |
| HMDB0241308 | Tridecanoylcarnitine | **0.541** | -0.229 | 0.153 | PC1: Mitochondrial lipid bioenergetics | 0.369 |
| HMDB0241362 | (12E)-10-Hydroxytetradec-12-enoylcarnitine | **0.874** | -0.043 | 0.119 | PC1: Mitochondrial lipid bioenergetics | 0.780 |
| HMDB0241694 | 5-Hydroxyoctanoylcarnitine | **0.875** | -0.103 | 0.123 | PC1: Mitochondrial lipid bioenergetics | 0.791 |
| HMDB0242462 | (2-Amino-3-hydroxyoctadecyl) dihydrogen phosphate | -0.086 | **0.891** | -0.081 | PC2: Sphingolipid stress signalling | 0.808 |
| HMDB0247790 | C18-Sphingosine 1-phosphate; D-erythro-Sphingosine-1-phosphate | -0.099 | **0.872** | -0.108 | PC2: Sphingolipid stress signalling | 0.782 |
| HMDB0250928 | Decylubiquinone | **0.707** | -0.231 | 0.334 | PC1: Mitochondrial lipid bioenergetics | 0.665 |
| HMDB0260560 | MG(0:0/18:3(9,11,15)-OH(13)/0:0) | **0.811** | -0.010 | 0.066 | PC1: Mitochondrial lipid bioenergetics | 0.662 |

**Component variance explained**

| **Component** | **Biological label** | **Initial eigenvalue** | **Unrotated variance (%)** | **Rotated variance (%)** | **Cumulative rotated variance (%)** |
| --- | --- | --- | --- | --- | --- |
| PC1 | Mitochondrial lipid bioenergetics | 8.485 | 42.427 | 39.559 | 39.559 |
| PC2 | Sphingolipid stress signalling | 3.221 | 16.103 | 17.688 | 57.248 |
| PC3 | Metabolic resilience | 2.310 | 11.548 | 12.831 | 70.079 |

Loadings are Varimax-rotated. For each metabolite, the loading corresponding to its principal component assignment is shown in bold. Principal assignment denotes the component with the largest absolute loading. Communality is the sum of squared loadings across the three retained components. The three rotated components explained 70.079% of the total variance.

**Abbreviations:** HMDB, Human Metabolome Database; PC, principal component; PCA, principal component analysis.

**ESF, Table S3. Composition and scoring of the eight non-PCA metabolic modules.**

| **Derived module** | **Constituent HMDB IDs / biomarkers** | **Constituent metabolites or biomarkers** | **Score definition** |
| --- | --- | --- | --- |
| Oxidative lipid damage + LOX | HMDB0060101; HMDB0260291 | 12-Hydroxyarachidonic acid; 5-(4-Carboxybutylperoxy)pentanoic acid | z(HMDB0060101) + z(HMDB0260291) |
| RAAS stress | HMDB0258895; HMDB0010357 | Tetrahydroaldosterone; Tetrahydroaldosterone-3-glucuronide | z(tetrahydroaldosterone) − z(tetrahydroaldosterone-3-glucuronide) |
| PUFA membrane remodelling | HMDB0010404; HMDB0244521; HMDB0265376; HMDB0267690 | LysoPC(22:6/0:0); 13,16-Docosadienoic acid; PA(PGJ2/20:5); PA(20:5-OH/i-15:0) | z(HMDB0010404) + z(HMDB0244521) + z(HMDB0265376) + z(HMDB0267690) |
| ANTIOXIDANT STATE | HMDB0061643; HMDB0245425; HMDB0255357;  apolipoprotein A1;  HDL-C;  albumin | 3-Carboxy-4-methyl-5-pentyl-2-furanpropanoic acid (CMPF); 2,3,4-Trihydroxybutanoic acid; 2-(20-Hydroxyicosa-5,14-dienoylamino)acetic acid; apolipoprotein A1; high-density lipoprotein cholesterol; albumin | z(CMPF) + z(2,3,4-trihydroxybutanoic acid) + z(2-(20-hydroxyicosa-5,14-dienoylamino)acetic acid) + z(apolipoprotein A1) + z(HDL-C) + z(albumin), followed by re-standardization. |
| Taurine | HMDB0000251 | Taurine | z-standardised abundance |
| Nonate | HMDB0011717 | Nonate | z-standardised abundance |
| Terpenoid metabolism | HMDB0035272; HMDB0032369; HMDB0039893; HMDB0035337 | p-Menthan-2-one; L-Menthyl acetoacetate; (1R,2R,4S)-p-Menthane-1,2,8-triol; Sterebin A | z(p-Menthan-2-one) − z(L-Menthyl acetoacetate) − z((1R,2R,4S)-p-Menthane-1,2,8-triol) − z(Sterebin A) |
| Alkamide | HMDB0030951 | (E,E)-2,4-Decadienoic isobutylamide | z-standardised abundance |

These eight non-PCA modules, together with PC1, PC2, and PC3, constitute the final set of 11 biologically informed metabolic modules. All z terms denote standardised values. For composite scores containing subtraction, the sign indicates the direction of contribution to the derived variable.

**Abbreviations:** HDL-C, high-density lipoprotein cholesterol; HMDB, Human Metabolome Database; LOX, lipoxygenase; PC, principal component; PCA, principal component analysis; PUFA, polyunsaturated fatty acid; RAAS, renin–angiotensin–aldosterone system; z, standardised value.

**ESF, Figure S1. Differential serum metabolite profiles across diagnostic comparisons.**

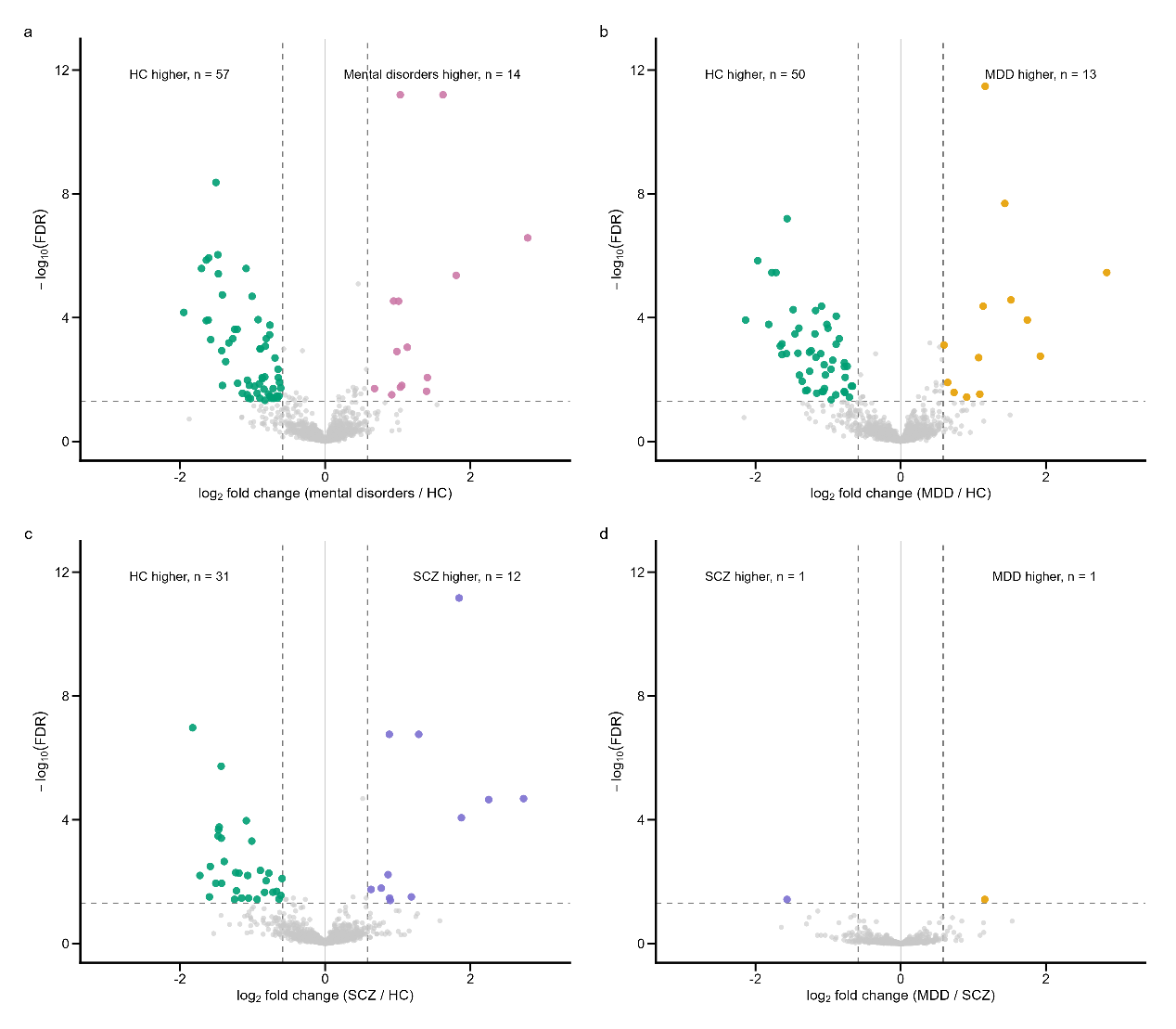

Volcano plots show differential serum metabolites for **(A)** pooled mental disorders versus healthy controls (HC), **(B)** major depressive disorder (MDD) versus HC, **(C)** schizophrenia (SCZ) versus HC, and **(D)** MDD versus SCZ. Each point represents one metabolite. Colored points indicate metabolites meeting the predefined statistical thresholds of false discovery rate (FDR) < 0.05 and absolute fold change ≥ 1.5, with colors denoting the group in which metabolite abundance was higher; gray points indicate non-significant metabolites. Vertical dashed lines indicate the fold-change threshold, and the horizontal dashed line indicates the FDR threshold. Numbers shown at the top of each panel indicate the metabolites meeting both statistical criteria in each direction. Identical x- and y-axis ranges are used across all panels to facilitate direct comparison of effect sizes and statistical evidence.

**Abbreviations**: FDR, false discovery rate.

**ESF, Figure S2. Discriminative performance of the 38-metabolite panel in the locked internal held-out test set.**

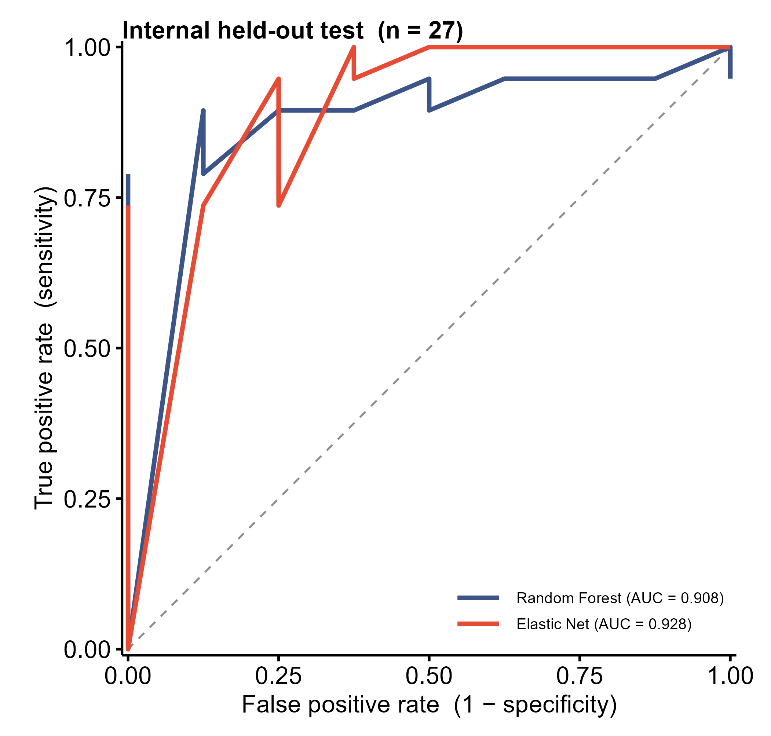

Receiver operating characteristic (ROC) curves show discrimination of healthy controls from pooled participants with major depressive disorder (MDD) or schizophrenia (SCZ) in the locked internal held-out test set (n = 27). Random-forest and elastic-net classifiers achieved AUCs of 0.908 and 0.928, respectively. Both classifiers correctly classified 22 of 27 participants, corresponding to an overall accuracy of 0.815. The diagonal dashed line indicates chance-level discrimination.

**ESF, Figure S3. OPLS-DA discrimination and permutation validation across alternative predictor combinations.**

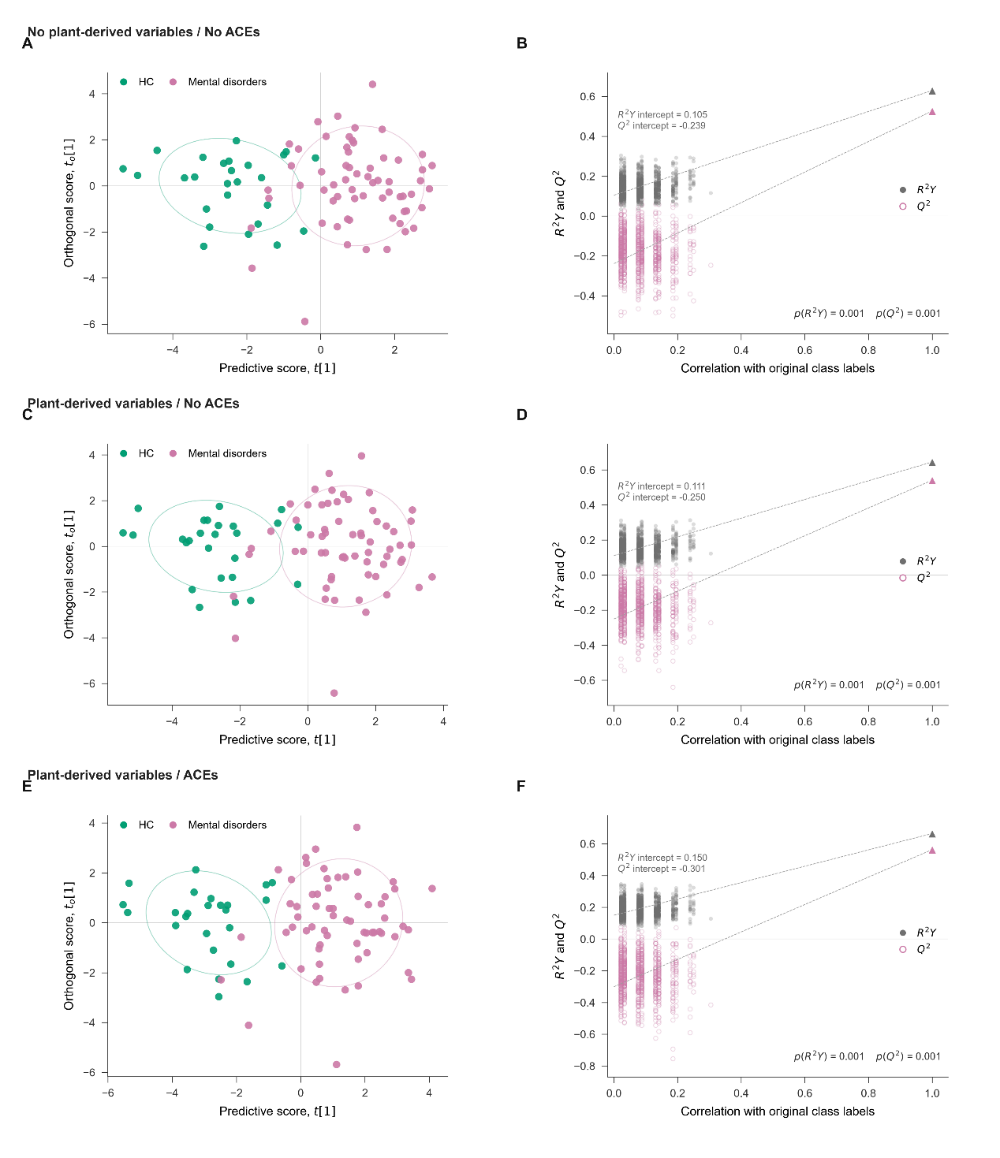

OPLS-DA models were evaluated using three predictor combinations complementary to the main Figure 5 model. **(A, B)** Model excluding both plant-derived variables and adverse childhood experience (ACE) variables. **(C, D)** Model including plant-derived variables but excluding ACE variables. **(E, F)** Model including both plant-derived variables and ACE variables. The plant-derived predictor set comprised terpenoid metabolism and alkamide. **(A, C, E)** OPLS-DA score plots showing separation between healthy controls (HC) and participants with mental disorders. Ellipses indicate 1.5-SD group covariance regions for visualization. **(B, D, F)** Permutation validation based on 1,000 permutations of the class labels. Permuted R²Y values are shown as filled gray circles and Q² values as open magenta circles; dashed lines indicate the corresponding regression lines, and triangles at a correlation of 1 represent the original models. Regression intercepts and empirical permutation *P* values are shown within each panel.

**Abbreviations:** HC, healthy controls; OPLS-DA, orthogonal partial least-squares discriminant analysis; R²Y, explained variation in the response; Q², cross-validated predictive ability.

**ESF, Figure S4. Variable importance and predictive performance across alternative OPLS-DA models.**

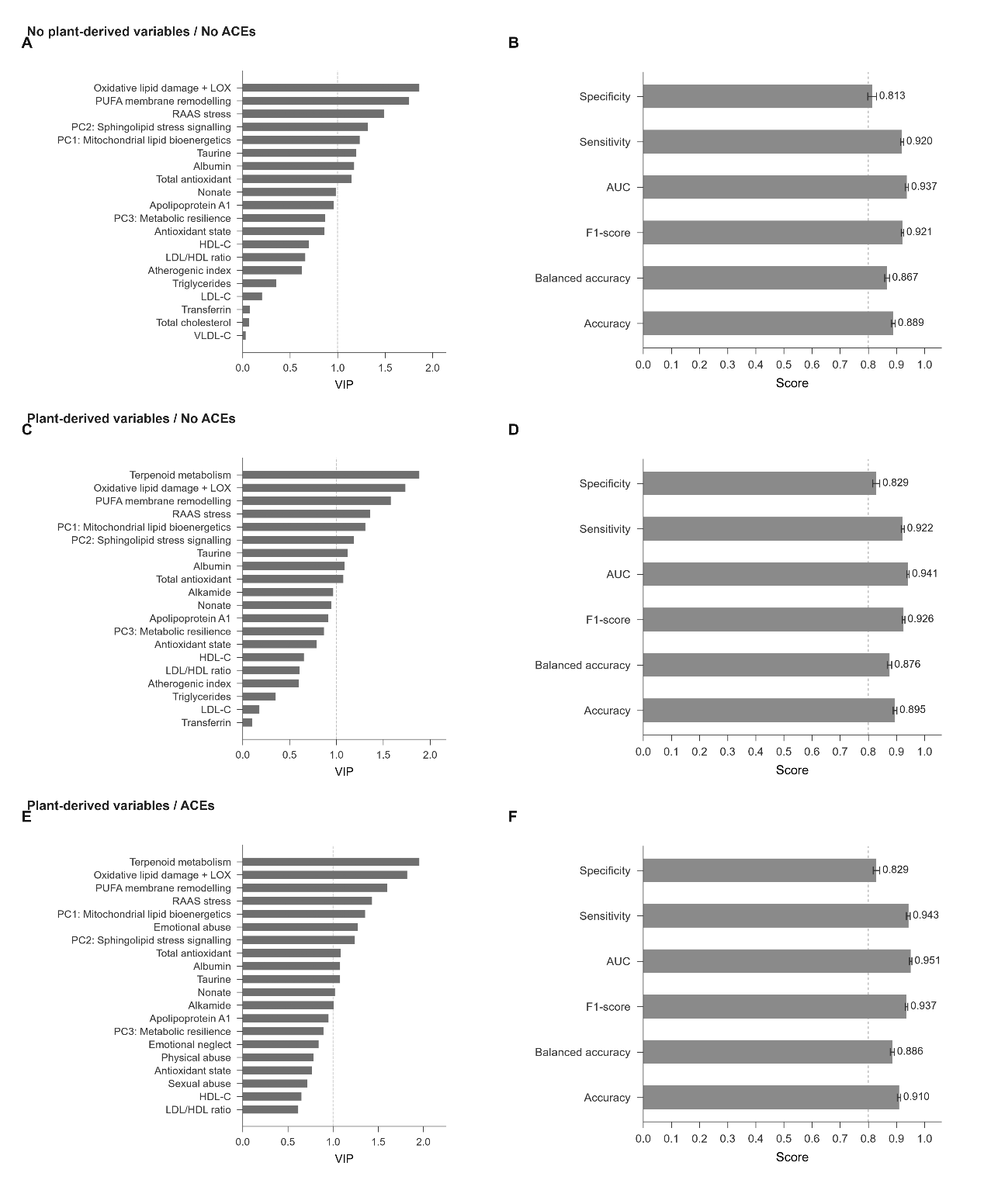

Variable importance in projection (VIP) rankings and nested cross-validation performance are shown for three alternative predictor configurations. (A, B) Model excluding both putatively diet-derived modules and childhood-adversity domains. (C, D) Model including the putatively diet-derived modules but excluding childhood-adversity domains. (E, F) Model including both putatively diet-derived modules and childhood-adversity domains. The putatively diet-derived modules comprised terpenoid metabolism and alkamide. (A, C, E) The 20 highest-ranking predictors according to VIP score are shown in descending order; the vertical dashed line indicates VIP = 1.0. (B, D, F) Predictive performance assessed by nested cross-validation, including specificity, sensitivity, area under the receiver operating characteristic curve (AUC), F1-score, balanced accuracy, and accuracy. Bars show point estimates, whiskers indicate 95% intervals where available, and the vertical dashed line denotes a reference value of 0.80.

**Abbreviations**: OPLS-DA, orthogonal partial least-squares discriminant analysis; VIP, variable importance in projection.
